# Safety and Immunogenicity of Nanocovax, a SARS-CoV-2 Recombinant Spike Protein Vaccine

**DOI:** 10.1101/2021.07.22.21260942

**Authors:** Thuy P. Nguyen, Quyet Do, Lan T. Phan, Duc V. Dinh, Hiep Khong, Luong V. Hoang, Thuong V. Nguyen, Hung N. Pham, Men V. Chu, Toan T. Nguyen, Tri M. Le, N.T. Tuyen, Thanh T. Dinh, Thuong V. Vo, Thao T. Vu, Quynh B.P. Nguyen, Vuong T. Phan, Luong V. Nguyen, Giang T. Nguyen, Phong M. Tran, Thuan D. Nghiem, Tien V. Tran, Tien G. Nguyen, Tuynh Q. Tran, Linh T. Nguyen, Anh T. Do, Dung D. Nguyen, Son A. Ho, Viet T. Nguyen, Dung T. Pham, Hieu B. Tran, Son T. Vu, Su X. Tran, Trung M. Do, Ton Tran, Thang M. Cao, Huy M. Dao, Thao T.T. Nguyen, Uyen Y Doan, Vy T.T. Le, Linh P. Tran, Ngoc M. Nguyen, Ngoc T. Nguyen, Hang T.T. Pham, Quan H. Nguyen, Hieu T. Nguyen, Hung T. Trinh, Hang L.K. Nguyen, Vinh T. Tran, Mai T.N. Tran, Truc T.T. Nguyen, Phat T. Ha, Hieu T. Huynh, Khanh D. Nguyen, Chung C. Doan, Si M. Do

## Abstract

**Background:** Nanocovax is a recombinant severe acute respiratory syndrome coronavirus 2 subunit vaccine composed of full-length prefusion stabilized recombinant SARS-CoV-2 spike glycoproteins (S-2P) and aluminum hydroxide adjuvant.

**Methods:** We conducted a dose-escalation, open label trial (phase 1) and a randomized, double-blind, placebo-controlled trial (phase 2) to evaluate the safety and immunogenicity of the Nanocovax vaccine (in 25 microgram (mcg), 50 mcg, and 75 mcg doses, aluminum hydroxide adjuvanted). In phase 1, 60 participants received two intramuscular injection of the vaccine following dose-escalation procedure. The primary outcomes were reactogenicity and laboratory tests to evaluate the vaccine safety. In phase 2 which involved in 560 healthy adults, the primary outcomes are vaccine safety; and anti-S IgG antibody response. Secondary outcomes were surrogate virus neutralization, wild-type SARS-CoV-2 neutralization, and T-cell responses by intracellular staining (ICS) for interferon gamma (IFNg). We performed primary analyses at day 35 and day 42.

**Results:** For phase 1 study, no serious adverse events (SAE) were observed for all 60 participants. Most adverse events (AE) were grade 1 and disappeared shortly after injection. For phase 2 study, after randomization, 480 participants were assigned to receive the vaccine with adjuvant, and 80 participants were assigned to receive placebo. Reactogenicity was absent or mild in the majority of participants and of short duration (mean, ≤3 days). Unsolicited adverse events were mild in most participants. There were no serious adverse events related to Nanocovax. Regarding the immunogenicity, Nanocovax induced robust anti-S antibody responses. There was no statistical difference in antibody responses among dose strengths on Day 42, in terms of anti S-IgG level and neutralizing antibody titer.

**Conclusions:** Up to 42 days, Nanocovax vaccine was safe, well tolerated and induced robust immune responses. We propose using Nanocovax 25 mcg for Phase 3 to evaluate the vaccine efficacy. (Research funded by Nanogen Pharmaceutical Biotechnology JSC., and the Ministry of Science and Technology of Vietnam; ClinicalTrials.gov number, NCT04683484.)

## 1. Introduction

Global pandemic coronavirus disease 2019 (Covid-19) is caused by the severe acute respiratory syndrome coronavirus 2 (SARS-CoV-2). As of July 2021, more than 177 million cases and over 4 million deaths due to Covid-19 have been reported worldwide^1^.

SARS-CoV-2 is a member betacoronavirus, named for its corona of spike (S) proteins protruding from the viral envelope^2,3^. SARS-CoV-2 S, a heavily glycosylated protein, is responsible for the attachment to angiotensin-converting enzyme (ACE2) which helps the virus entry to host cells in human and animals^4^. SARS-CoV-2 S glycoprotein is the antigen of choice for Covid-19 vaccine development due to its highly antigenic property^5^.

Nanocovax is a subunit vaccine, developed and manufactured at Nanogen Pharmaceutical Biotechnology JSC., containing full-length prefusion stabilized recombinant SARS-CoV-2 S glycoproteins and aluminum hydroxide adjuvant. In rodent and monkey models, Nanocovax induced high levels of anti-S antibody (Ab). Neutralizing antibody titers were evaluated by microneutralization on Wuhan strain and surrogate virus neutralization test. Importantly, Nanocovax conferred a remarkable protection against SARS-CoV-2 infection in hamster challenge model^6^.

Here we report the findings of the phase 1 and 2 trials started in December 2020 and February 2021 respectively, to evaluate the safety and immunogenicity of 25 mcg, 50 mcg and 75mcg dose strengths of recombinant SARS-CoV-2 S glycoprotein with aluminum adjuvant (0.5 mg/dose) in healthy adults of at least 18 years of age.

## 2. Method

### 2.1. Trial design and oversight

Phase 1 trial was conducted at the Military Medical University, Ha Noi, Vietnam. This was an open-labeled, dose-escalation study with the emphasis on the vaccine safety (figure S1). Eligible participants were healthy men and nonpregnant women, 18 to 50 years of age with body-mass index (BMI) of 18 to 27 kg/m^2^ (table S2). In this phase, 60 participants were allocated into 2:2:2 ratio of 25 μg, 50 μg and 75 μg dose groups, respectively. For safety measure, the first 3 participant of 25 μg dose group were vaccinated and monitored for 72 hours. After no SAE were observed, all remaining participants in this group plus 3 participants in 50 μg dose group would be vaccinated and monitored for 72 hours. If no SAE were observed among 3 participants of 50 dose group, the remaining participant in this group plus 3 participants in 75 μg dose group would be vaccinated. If no SAE were observed among 3 participants in 75 μg dose group, the remaining participants in this group will be vaccinated. All participants received 2 intramuscular injections of the vaccine into the deltoid on day 0 and day 28. Sample size of phase 1 was not based on formal statistical power calculation but on the range of 30 – 150 recommended in Article 10 of Appendix 10/2020/TT-BYT by Vietnam Ministry of Health.

Phase 2 trial was conducted at two sites: Military Medical Academy, Ha Noi and the Pasteur Institute at Ho Chi Minh city, Vietnam. This was a randomized, double-blind, placebo-controlled study (figure 1). Eligible participants were healthy men and nonpregnant women, at least 18 years of age with BMI of 17 to 35. They were stratified into 3 age groups: from full 18 to full 45 years old, from 46 to 60 years old, and over 60 years old (table S2). The sample size of phase 2 was calculated based on the estimated probability of observing an adverse event. Accordingly, a total of 560 participants will be randomly assigned to 4 groups, into 2:2:2:1 ratio for 25 μg, 50 μg, 75 μg, and placebo, respectively. In details, 480 volunteers would receive the vaccine (160 volunteers receiving 25 µg dose; 160 volunteers receiving 50 µg dose and 160 volunteers receiving dose of 75 µg dose) and 80 volunteers would receive the placebo (aluminum adjuvant only). All participants received 2 intramuscular injections of the vaccine or the placebo into the deltoid on day 0 and day 28. Trial staffs responsible for the vaccine preparation and administration, and participants were unaware of vaccine assignment. Randomization lists, using block randomization stratified by study group and study site, were generated by the study statistician. Computer randomization was done with full allocation concealment within the secure web platform used for the study electronic case report form (service provided by Medprove company).

**Figure 1.**
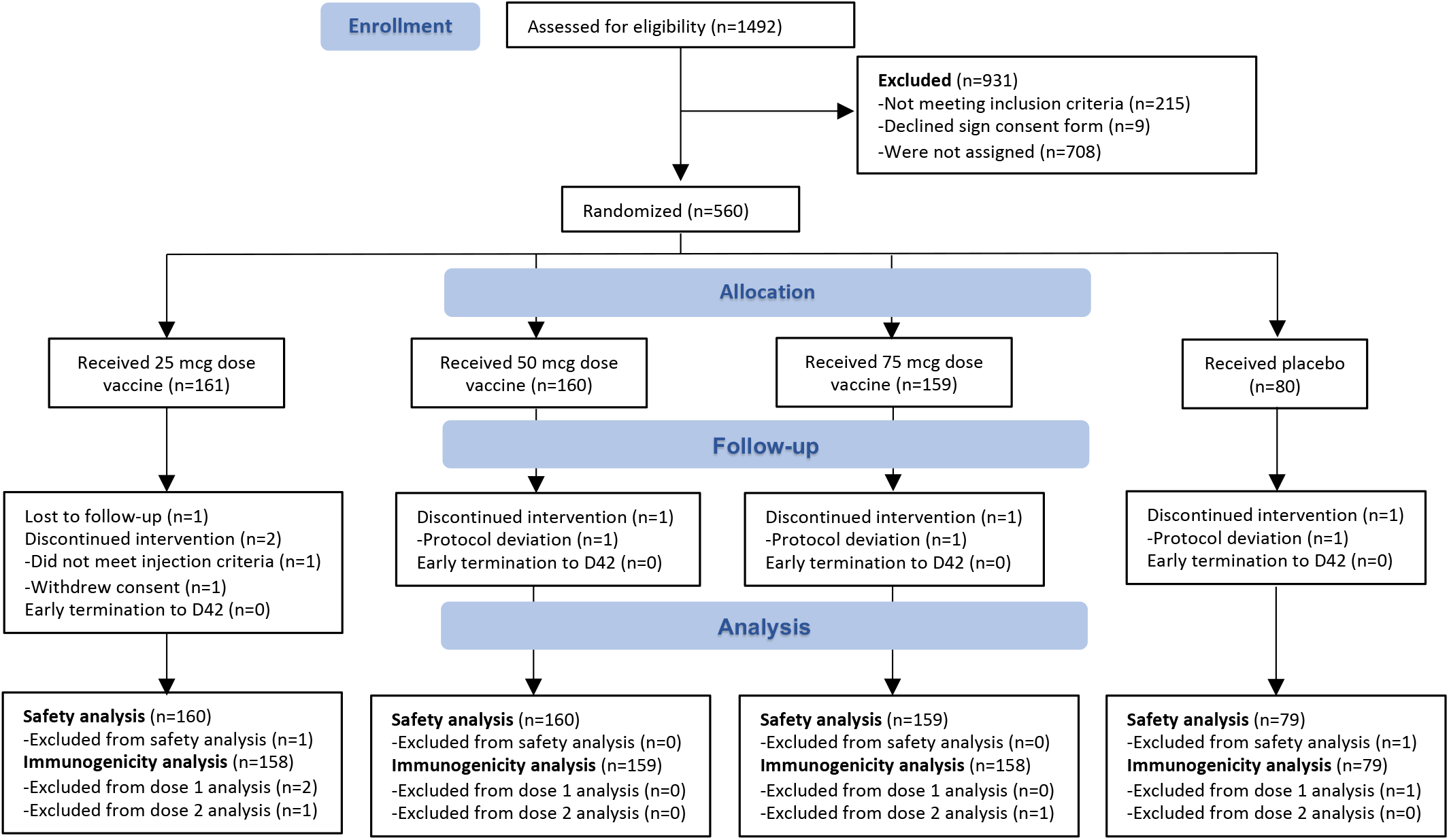
Screening and randomization of participants in phase 2.

All participants were screened by their medical history, clinical and biological examinations, sampling and laboratory tests (complete blood count, biochemistry, urine analysis, testing pregnancy and diagnostic imaging). Participants with a history of Covid-19 or positive results for SARS-CoV-2 at screening period confirmed by real-time reverse transcriptase polymerase-chain-reaction (RT-PCR) were excluded from the trials. All participants provided written consent before being enrolled into the trial.

The trials were designed and funded by Nanogen Pharmaceutical Biotechnology JSC and the Ministry of Science and Technology (MOST) of Vietnam. The trial protocol was approved by the Ethics Committee/Protocol Review Board of the Ministry of Health (Vietnam) and was performed in accordance with the ICH-GCP good clinical practice guidelines, with an ethical policy consistent with the “Declaration of Helsinki” and applicable Vietnamese laws and regulations. The authors take responsibility for the integrity of the data and the fidelity of the trial to the protocol.

### 2.2. Trial vaccine, adjuvant, and placebo

The recombinant SARS-CoV-2 spike (S) glycoprotein in Nanocovax were constructed with 682-QQAQ-685 mutations for protease resistance, and two proline substitutions (K986P and V987P) for stabilized prefusion conformation. The production of the full-length (including the transmembrane domain) recombinant S protein was optimized in the established Chinese Hamster Ovary (CHO) cell-expression system. Clinical grade aluminum hydroxide was manufactured by Croda (Denmark). Recombinant SARS-CoV-2 S protein were absorbed to aluminum adjuvant in mild shaking condition for 18 hours at 2°C to 8°C. Placebo was sterile 0.05% aluminum.

### 2.3. Safety assessments

In phase 1, the onsite safety follow-up time after was 72 hours after 1^st^ injection and 24 hours after the 2^nd^ injection. Participants would return to the study site for follow-up visits at scheduled timepoints (table S1). In phase 2, the onsite safety follow-up time was 60 minutes after each vaccination. Follow-up visits to evaluate safety were scheduled on days 28, 35, 42, 90, 180 after vaccination (table 1). Participants were observed for 60 minutes after each vaccination for assessment of reactogenicity. In both phases, participants received instruction for self-monitoring and reporting adverse events during 7 days after each vaccination, as facilitated by the use of a diary with predefined reactogenicity. Predefined local (injection site) reactogenicity included pain, tenderness, erythema, and swelling. Predefined systemic reactogenicity included fever, nausea or vomiting, headache, fatigue, malaise, myalgia, and arthralgia. Vaccination pause rules were in place to monitor participants’ safety (Supplementary appendix).

**Table 1.**
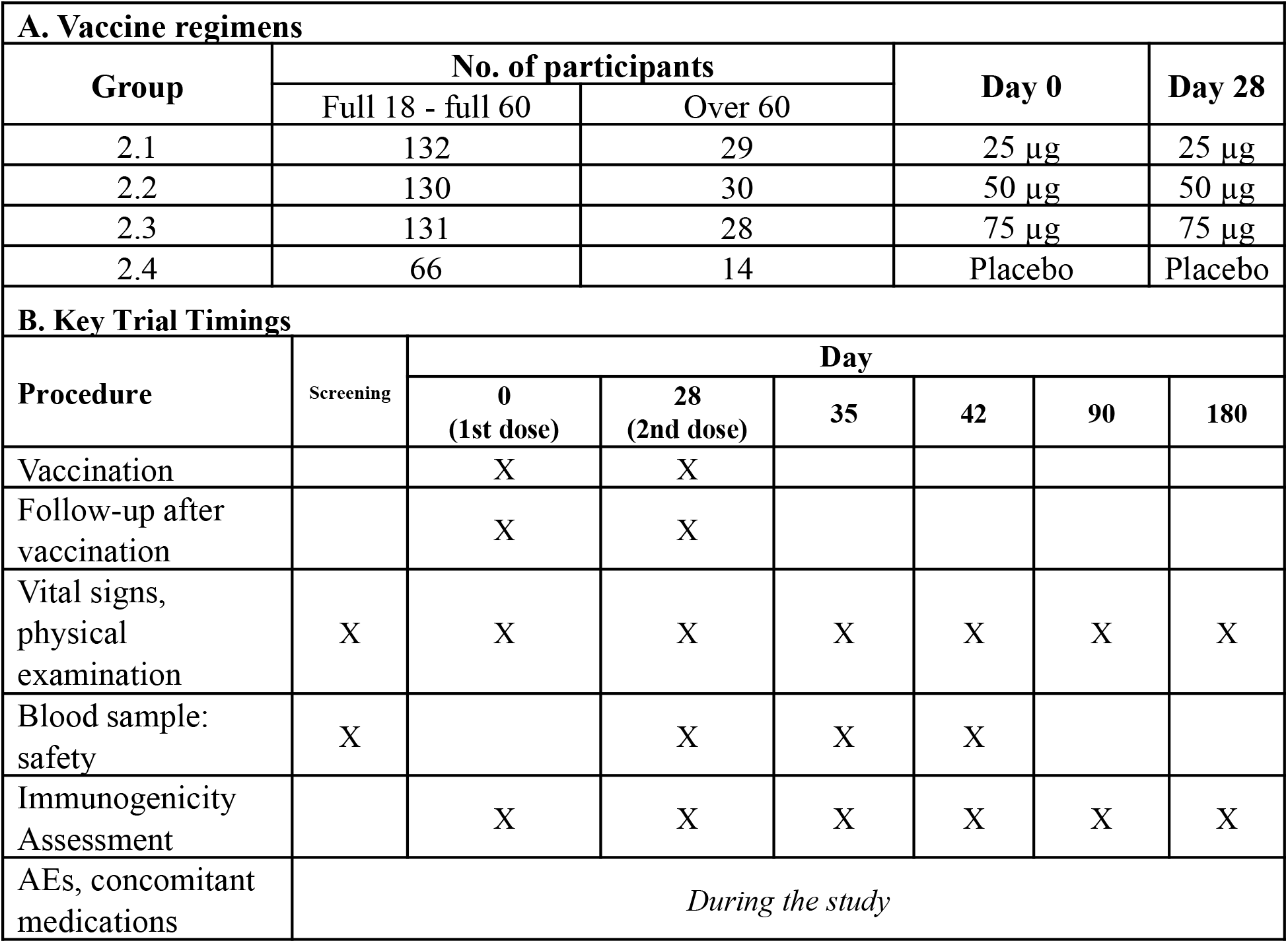
Vaccine regimens and key trial timings of phase 2. Shown are the planned schema and associated vaccine regimens administered in the trial (panel A), along with timing of the key safety and immunogenicity assessments (panel B).

**Table 2.** Demographic characteristics of the participants in the phase 2 trial at enrollment.

| Characteristics | Low dose<br>(25 mcg) | Medium dose<br>(50 mcg) | High dose<br>(75 mcg) | Placebo | Total |
| --- | --- | --- | --- | --- | --- |
| **Group** | **2.1** | **2.2** | **2.3** | **2.4** |  |
| Number (n) | 161 | 160 | 159 | 80 | 560 |
| **Age (in year)** |  |  |  |  |  |
| Mean ± SD | 43.7 | 44.4 | 44.9 | 42.6 | 44.1 |
| Range (min - max) | 19 - 76 | 20 - 77 | 19 - 77 | 18 - 73 | 18 - 73 |
| **Age group – n (%)** |  |  |  |  |  |
| From 18 to 45 years old | 85 (52.8%) | 85 (53.1%) | 83 (52.2%) | 44 (55.0%) | 297 (53.0%) |
| From 46 to 60 years old | 47 (29.2%) | 45 (28.1%) | 48 (30.2%) | 22 (27.5%) | 162 (28.9%) |
| Over 60 years old | 29 (18.0%) | 30 (18.8%) | 28 (17.6%) | 14 (17.5%) | 101 (18.0%) |
| **Sex – n (%)** |  |  |  |  |  |
| Male | 69 (42.9%) | 75 (46.9%) | 63 (39.6%) | 35 (43.8%) | 242 (43.2%) |
| Female | 92 (57.1%) | 85 (53.1%) | 96 (60.4%) | 45 (56.3%) | 318 (56.8%) |
| **Ethnicity – n (%)** |  |  |  |  |  |
| Kinh | 99.4 | 98.1 | 98.7 | 100 | 98.9 |
| Other | 0.6 | 1.9 | 1.3 | 0 | 1.1 |
| **Body Mass Index**<br>**(BMI)\*** |  |  |  |  |  |
| Number (n) | 161 | 160 | 159 | 80 | 560 |
| Mean ± SD | 23.1 ± 3 | 23.1 ± 3.4 | 22.9 ± 2.9 | 22.7 ± 2.8 | 23 ± 3.1 |
| Range (min - max) | 17 - 32 | 16 - 33 | 17 - 31 | 17 - 28 | 16 - 33 |
| **BMI ≥ 30** |  |  |  |  |  |
| n | 5 | 7 | 4 | 0 | 16 |
| % | 3.11 | 4.38 | 2.52 | 0 | 2.86 |

\* The body-mass index is the weight in kilograms divided by the square of the height in meters.

The primary safety outcomes were the number and percentage of participants with solicited local and systemic adverse events occurred within 7 days after vaccination and laboratory results (serum chemistry and hematology) at days 0, 7, 28, 35 according to FDA toxicity scoring^7^. Secondary safety outcomes were occurrence rate and severity rating of unsolicited AE/SAE through day 56 and laboratory results at days 42 and 56 (phase 2 and 1, respectively).

AEs/SAEs were recorded and evaluated basing on the Common Terminology Criteria for Adverse Events 5.0 (CTCAE v5.0) and Guidelines for assessing toxicity in healthy volunteers in FDA’s Preventive Vaccine Clinical Trial Study^7,8^. The procedures for recording and evaluating take place continuously from the time of using the first dose to the end of the last visit in each research volunteer. Adverse events were assessed in terms of severity score (mild, moderate, severe, life-threatening, or fatal), and relatedness to the vaccine. Vital sign measurements were assessed according to FDA toxicity scoring after vaccination. In addition, participants underwent nasopharyngeal swab testing for SARS-CoV-2 on screening day (before 1^st^ vaccination), day 28 (before 2^nd^ vaccination) and any time that they developed symptoms of possible SARS-CoV-2 infection.

### 2.4. Immunogenicity assessments

The primary outcome was anti-S IgG responses to Nanocovax evaluated by chemiluminescence immunoassay (CLIA). Secondary outcomes were neutralizing antibody titer evaluated by 50 percent plaque reduction neutralization test (PRNT_50_) on Wuhan strain and UK variant (B.1.1.7), neutralizing activity evaluated by competitive enzyme-linked immunosorbent assay (ELISA) based surrogate virus neutralization test (sVNT), and T cell response by intracellular cytokine-staining (ICS). Details of these assays are provided in the supplementary appendix.

### 2.5. Statistical analysis

#### 2.5.1. Safety analysis

Local and systemic adverse events occurred during 7 days after each injection were documented (number, percentage) by group (vaccine and placebo) and analyzed to find factors (if any) correlated/associated with the severity of AE.

Serious adverse events were classified according to CTCAE 5.0 and statistically described according to the number and percentage of each research group. If an adverse event occurs more than once, analysis is based on only the most severe occurrence and the cause of the event. In addition, all serious adverse events will be summarized separately.

The results were statistically described as number of and percentage of volunteers having abnormal results or have results changed overtime, by group, compared to baseline values. Values higher than normal ranges/thresholds will be assessed for clinical significance by researchers.

AE/SAEs were statistically analyzed the difference in percentage cumulative with 95% confidence interval of the Clopper-Pearson method (Clopper-Pearson 95% CIs) between vaccine dose groups.

#### 2.5.2. Immune response analysis

The geometric anti-S IgG antibody and neutralizing ability of SARS-CoV-2 antibody in the sera at the screening stage is considered as baseline values. Geometric mean concentration (GMC) of anti-S IgG and geometric mean titers (GMT) of neutralizing antibody after vaccination are evaluated at defined time points in table 1.

Data were analyzed base on two-sided test with 95% confidence in the t-distribution function: anti-S IgG titer should increase at least 4 times (Clopper-Pearson 95% CIs) as compared to baseline, at defined time points post vaccination, in each dose group.

## 3. Result

### 3.1. Trial population

The phase 1 trial was started on December 17, 2020. 60 participants underwent randomization into 3 groups of doses: 20 received 25 mcg (group 1.1), 20 received 50 mcg (group 1.2), and 20 received 75 mcg (group 1.3). There was no placebo group for this stage of study, and the primary assessment was the vaccine safety.

For phase 2, the study was started on February 26, 2021. 560 participants were recruited and randomly assigned into groups of different doses: 161 received 25 mcg doses of Nanocovax (group 2.1), 160 received 50 mcg doses of Nanocovax (group 2.2), 159 received 75 mcg doses of Nanocovax (group 2.3), and 80 received placebo (group 2.4). Participants were stratified into 2 age groups: from over 18 to below 60 years old and over 60 years old. Demographic characteristics of participants in phase 1 and 2 are shown in table 1 and table S1.

### 3.2. Safety outcomes

In phase 1, no serious adverse events were observed, and vaccination pause rules were not implemented. Overall reactogenicity was largely absent or mild, and second vaccinations were neither withheld nor delayed due to reactogenicity. The percentage of participants in each vaccine group (groups 1.1, 1.2, and 1.3) with solicited adverse events according to the maximum FDA toxicity grade (grade 1: mild, grade 2: moderate, grade 3: severe, grade 4: life-threatening) during the period of this study was presented in figure S2. These preliminary results showed that Nanocovax were safe at all dose strength: 25 mcg, 50 mcg, and 75 mcg.

In phase 2, among 560 participants, there were 554 participants receiving full 2 dose of vaccine or placebo (figure 1). 6 withdrew from the study after the 2^nd^ visit, before getting the boosting dose. Local AE were 37.7% after 1st vaccination and 35.6% after 2nd vaccination. Local pain were 32.6% (181/556) after 1st injection and 32.1% (178/554) after 2nd injection. Systemic AEs were 27.4% after 1^st^ injection and 21.8% after 2^nd^ injection. Most common systemic AEs were fatigue (16.9%/13%), headache (13.3%/8.3%) and fever (3.8%/2.4%) after 1^st^/2^nd^ injections. After 1^st^ injection, local and systemic AE of grade 3 or 4 were not observed in group 2.1, 1 case in group 2.2 (0.6%), 1 case in group 2.3 (0.6%) and 1 case in group 2.4 (1.3%). After 2^nd^ injection, local and systemic AE of grades 3 or 4 were 1 in each vaccine group (0.6%) and placebo (1.3%) (figure 2).

**Figure 2.**
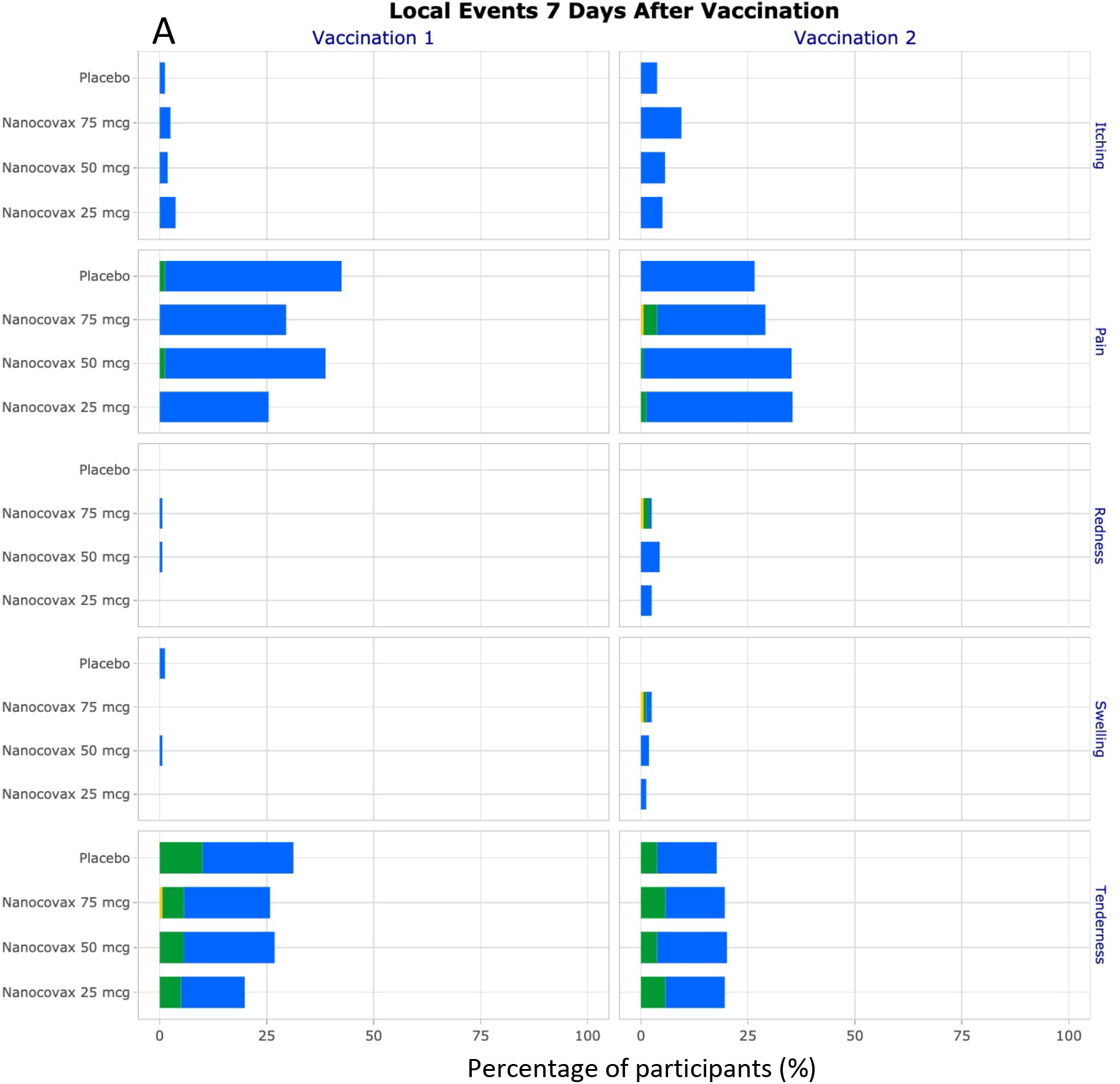
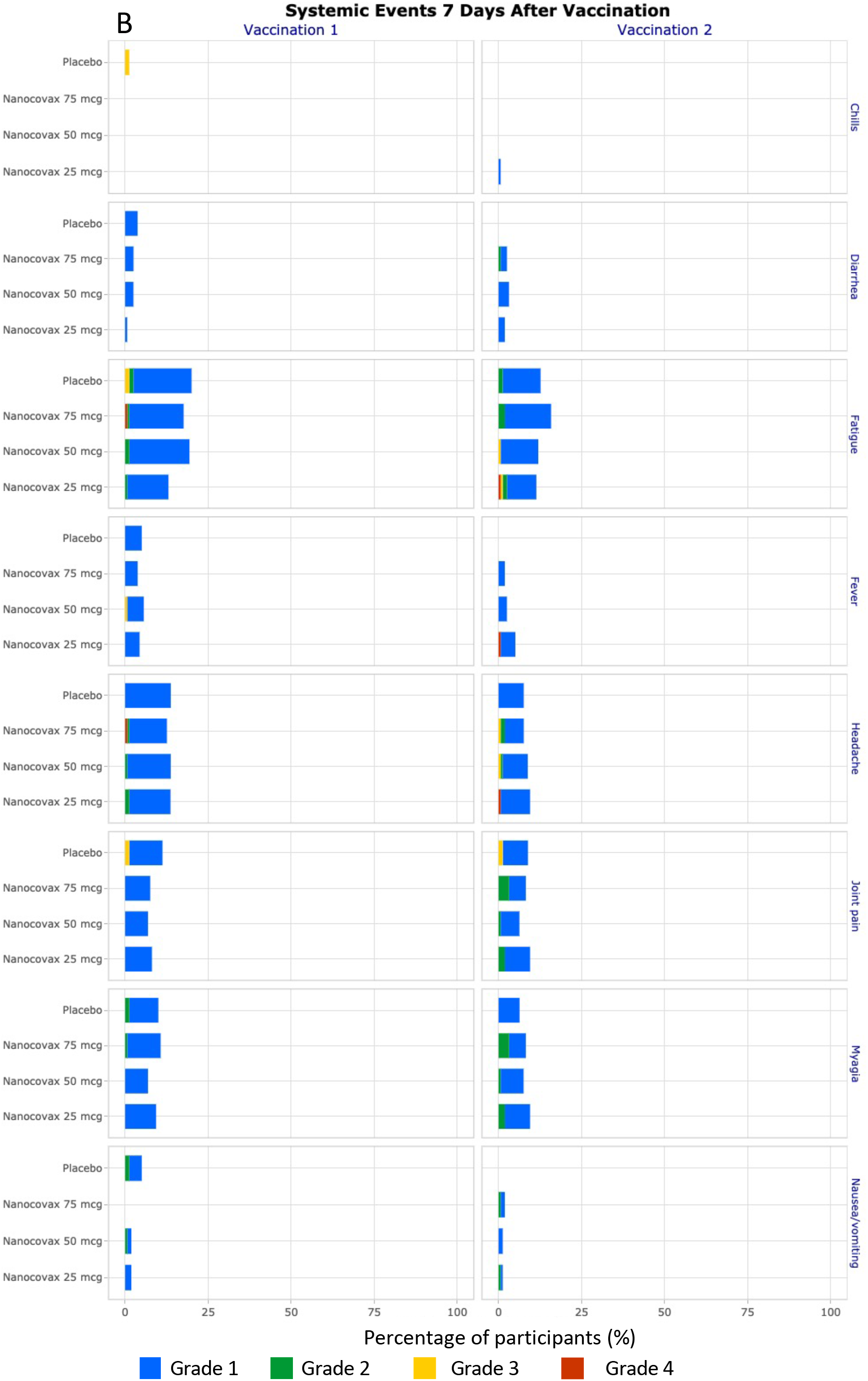
Solicited local adverse event (A) and systemic adverse events (B) within 7 days after vaccination in phase 2.

The incidence of unsolicited AEs in were similar in vaccine groups and the placebo (appendix 7). In details, unsolicited AE incidence of groups 2.1 to 2.4 were 30.4%, 27.5%, 23.3% and 33.8%, respectively. Two cases of grade 4 AE were back pain and dizziness. There were five AE of grade 3: 1 case of sepsis, 1 case of back pain, 1 case of spondylolisthesis, 1 case of sore throat, 1 case of high blood pressure. The most frequently reported adverse events were sore throat 27 (4.8%) and coughing 11 (2%). The most laboratory-related AEs were hyperglycemia 13 cases (2.3%), leukocytosis: 8 cases (1.4%), the most vital events related to hypothermia with 12 cases (2.1%). In similar to phase 1, vaccination pause rules were not implemented in phase 2. Four SAE were determined unrelated to Nanocovax, including 1 case of angina (history of stent graft), 1 case of fever (determined to be sepsis), 1 case of abscess at axillary lymph nodes occurred on the unvaccinated arm and 1 case of personal injury. One case SAE grade 1 of anaphylaxis was undetermined to be related to vaccine or not because the symptoms were not clear (table S3).

The percentage of participants in each vaccine group (groups 2.1, 2.2, 2.3, and 2.4) with adverse events according to the maximum FDA toxicity grade during the 7 days after each vaccination were plotted for solicited local and systemic adverse events (figure 2). There were no grade 4 events. The most adverse event was pain at the injection site, mild pain (grade 1) accounted for 23.4% (130/556) after 1st injection and accounted for 26.9% (149/554) after 2nd injection. Moderate pain (grade 2), was uncommon, accounted for 0.4% (2/556) after the 1st injection and 1.1% (6/554) after the 2nd injection. Mild sensitivity at the injection site accounted for 14.7 % (82/556) after 1st injection and 14.3% (79/554) after 2nd injection. Moderate sensitivity accounted for 4% (22/556) after injection 1 and 3.8% (21/554) after injection 2. The severe event (grade 3) was “pain at injection site” appearing in 1 subject after 2nd injection, accounting for 0.2% (1/554) and “redness at injection site” occurring in 2 subjects (2/554) after 2nd injection. The events mostly occurred within 2 days after vaccination, gradually decreased and disappeared within 7 days after vaccination. Some subjects felt fatigue after injection, accounting for 9.4% (52/556) after 1st injection and 8.3% (46/554) after 2nd injection. Fatigue decreased gradually day by day, disappeared at about 7 days after injection. The event “fever” occurred in relatively few subjects, mild fever accounted for 0.2% (1/556) after 1st injection and 1.1% (6/554) after 2nd injection. There was 1 subject with high fever (grade 3) accounted for 0.2% (1/556), occurred from day 1 to day 5 after 2nd injection. In the phase 2 study, the proportion of subjects with any “unsolicited” AE after injection with vaccine and placebo was 157/560 (28%). The rates of any AE in the study groups were similar between the 25 mcg (30.4%), 50 mcg (27.5%), 75 mcg (23.3%) and placebo (33.8%) groups. Most of “unsolicited” AEs were mild to moderate. When comparing the overall incidence of unsolicited AEs between the vaccine and placebo groups, the rates were similar in the two groups, with a ratio of 130/480 (27.1%) in the vaccinated group compared to the ratio of 27/80 (33.8%) in the placebo group.

Laboratory abnormalities in phase 2 included increased white blood cell in 3 participants, (0.6%), increased neutrophil in 2 participants (0.4%), elevated ALT (grade 2) in 3 participants (0.6%), and elevated AST (grade 2) in 2 participants (0.4%). The other biochemistry and hematology parameters such as red blood cell (RBC), hemoglobin (HGB), creatinine, bilirubin, prothrombin time (PT) fluctuated within normal limits (appendix 7).

### 3.3. Immunogenicity outcomes

Anti-S IgG antibody in the serum was quantified by CLIA. Geometric mean concentration (GMC) of anti-S IgG (U/ml) was reported. Before the first injection, the GMC values of the 4 groups were all below the lower limit of detection. Anti-S IgG GMC of 3 vaccine groups increased sharply after 2nd injection, on day 35 and day 42 (figure 3). On day 35, GMC of group 2.1, 2.2 and 2.3 were 6.78 U/ml (95% CI: [5.09-9.03]), 9.38 U/ml [6.99 – 12.58], and 13.04 U/ml [9.46 – 17.98] respectively. GMC of group 2.3 was statistically higher than that of group 2.1 but not group 2.2. GMC of the pairs (2.1 vs 2.2) and (2.2 vs. 2.3) were not statistically different. On day 42, GMC of group 2.1, 2.2 and 2.3 increased sharply: 60.48 U/ml [51.12 - 71.55]; 49.11 U/ml [41.26 - 58.46] and 57.18 U/ml [48.4-67.5], respectively. Meanwhile, the GMC of the placebo group at days 35, and 42 were 0.29 U/ml [0.25 – 0.33], and 0.29 U/ml [0.25 – 0.32], respectively.

**Figure 3.**
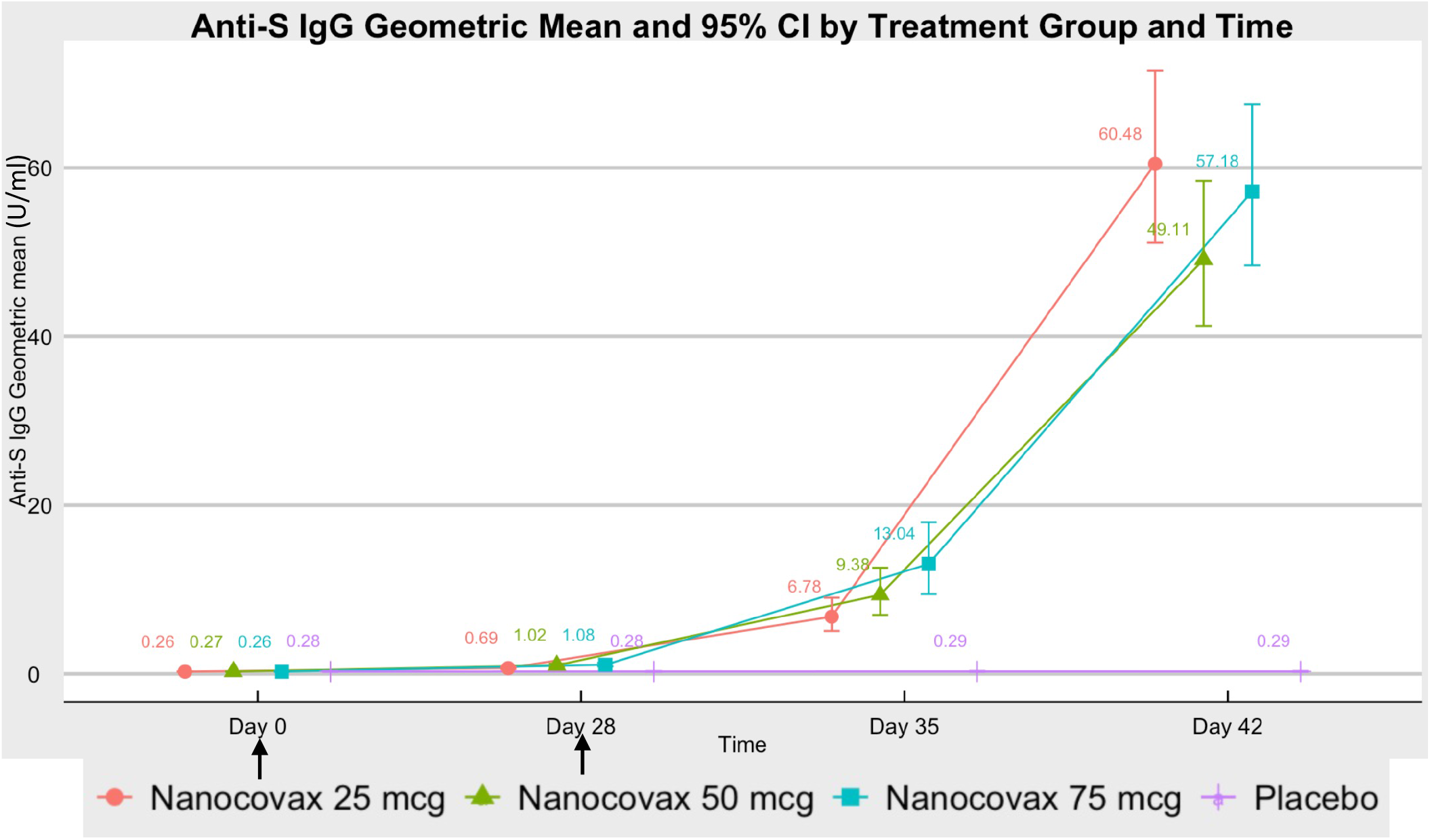
Anti-S IgG antibody responses of vaccine and placebo groups, expressed in geometric mean concentration. Arrows indicate days of vaccination. I bar represents 95% confident intervals (95% CI).

Geometric mean fold rise (GMFR) of anti-S IgG was defined as the fold increase in GMC of a given timepoint compared to baseline GMC value of the same group on day 0. GMFR of 2.1, 2.2 and 2.3 groups on day 35 were 25.7 [19.3 – 34.1], 34.7 [25.7 – 46.9], and 49.8 [36.0 – 68.9], respectively. At day 42, the GMFR of 2.1, 2.2 and 2.3 groups were 229.0 [193.2-271.4], 181.8 [152.0 – 217.4] and 218.3 [185.0 – 257.7]. Meanwhile, the GMFR of the placebo group (2.4) on days 35 and 42 were 1.05 and 1.04 respectively (figure S4).

The seroconversion rate was defined as GMFR > 4. Based on the GMFR of anti-S IgG, the seroconversion rates of 2.1, 2.2 and 2.3 group on day 35 were 84%, 84% and 85%, respectively. On day 42, the seroconversion rate of 2.1, 2.2 and 2.3 groups were 100%, 99% and 100%, respectively (Figure S5).

Surrogate virus neutralization test (sVNT) results, expressed as inhibition percentage (%), were considered positive if they were higher than cut-off value of 30%^9^. On day 35, the mean sVNT of groups 2.1 to 2.4 were 58.5% [54.1 – 63.0], 63.8% [59.1 – 68.5], 70.2% [65.8 – 74.5], 11.1% [9.3 – 12.7), respectively. Accordingly, the proportions of participants positive for sVNT in groups 2.1 to 2.4 were 80.4%, 82.4%, 86.7% and 1.3%, respectively. On day 42, the mean sVNT of group 2.1 to 2.4 were 87.5% [85.5 – 89.5], 86.4% [84.08 – 88.65], 87.1% [85.06 – 89.16], and 10.8% [8.9 – 12.67], respectively. Accordingly, the proportions of participants positive for SVN in groups 2.1 to 2.4 were 100%, 100%, 99.4% and 1.3%, respectively (figure 4A).

**Figure 4.**
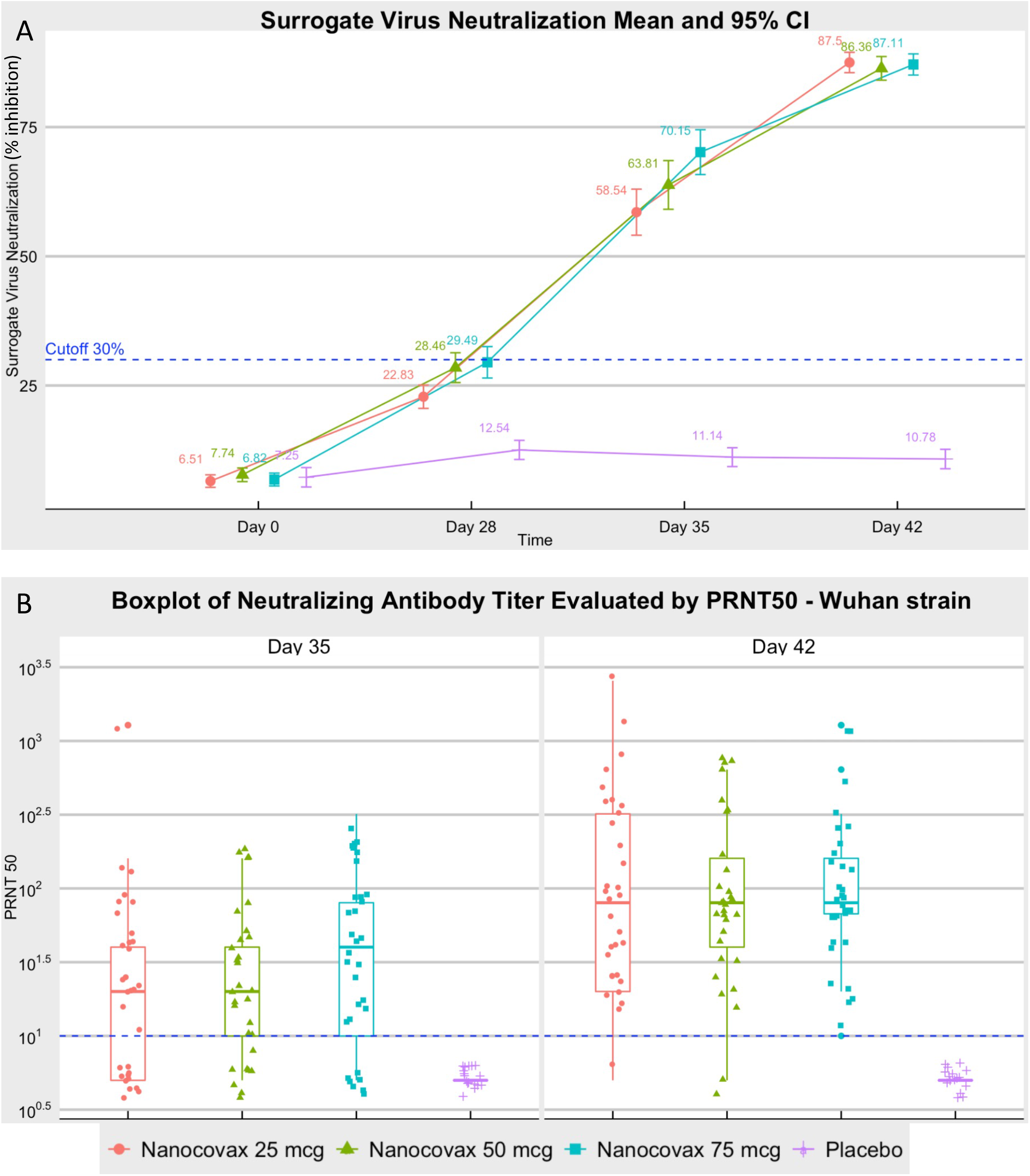
Neutralizing antibody responses. A) Surrogate virus neutralization was performed on all serum samples. Sera of day 0 were collected before 1^st^ injection. B) PRNT_50_ on the original Wuhan strain was performed on 112 randomly selected serum samples including vaccine groups (n=32 each) and placebo (n=16). Results expressed as geometric mean with 95% confidence interval. Dashed line indicates lower limit of detection. Each data point represents a sample. I bar represents 95% CI.

Neutralizing antibody titers were evaluated by plaque reduction neutralization test with inhibitory dilution greater than 50% (PRNT_50_). 112 serum samples of groups 2.1 to 2.4 were randomly selected for PRNT_50_ on Wuhan strain and UK variant. The results were expressed as geometric mean titer (GMT). On day 35, GMT of groups 2.1 to 2.3 were 20.9 [12.8 - 34.1], 22.5 [14.5 – 34.7] and 33.6 [20.9 – 54.1]. GMT of group 2.3 was statistically higher than that of group 2.1. However, no statistical difference was found between the pairs (2.1 vs. 2.2) and (2.2 vs. 2.3). On day 42, GMT of vaccine groups were 89.2 [52.2 – 152.3], 80 [50.8 – 125.9] and 95.1 [63.1 – 143.6], almost 3-time increase, compared to day 35. Meanwhile, serum samples of the placebo group at days 35 and 42 did not show neutralizing activity (figure 4B). As a matter of interest, a small subset of 112 serum samples were randomly selected to test neutralizing activity against UK variant (B.1.1.7, also known as the alpha variant). As expected, neutralizing titer found on UK variant was similar to that of Wuhan strain (figure S7).

T cell responses of 84 randomly selected participants (28 for each vaccine group and 14 for placebo group) were undetectable (data not shown). This was likely due to the nature of aluminum adjuvant which has been well established for Th2 response induction.

## 4. Discussion

The results of these phase 1 and phase 2 studies demonstrated an excellent safety profile of Nanocovax, regardless of dose strengths. Most adverse events and serious adverse events were grade 1 which disappeared within 48 hours after injection. Compared to similar studies of other approved vaccines, the vaccine may have the least reactogenicity^10–16^.

The vaccine was found to elicit high level of anti-S IgG which closely correlated with neutralizing antibody levels (figure S8). Importantly, PRNT results showed that the vaccine, regardless of dose strength, was effective against both original Wuhan strain and UK variant (B.1.1.7) (figure 4B and figure S7). Cellular immune response, evaluated by ICS for IFNg, was not observable. However, undetectable IFNg signal, a marker of Th1 response, does not guarantee the absence T cell response. In fact, it was likely due to the Th2 promoting nature of aluminum adjuvant^17^. Accordingly, T cell responses will be reevaluated in a small subset of participants in phase 3 with the addition of Th2 cytokines. The Th2 responses, if detected in phase 3, may raise a theoretical concern for vaccine-associated enhanced respiratory disease (ERD)^18,19^. This concern has been partially addressed with the aforementioned SARS-CoV-2 challenge on hamster model^6^. We will further evaluate the risk of ERD in phase 3 trial.

Although the efficacy of Nanocovax remains to be seen in a phase 3 trial, accumulated evidences have correlated the immunogenicity, particularly the neutralizing antibody level with the protection against Covid-19. Khoury and Cromer et al. provided a predictive model of efficacy by comparing the neutralizing antibody levels elicited by different vaccines to those of convalescent samples^20^. Their model suggested that the neutralizing level was highly predictive of immune protection against SARS-CoV-2 infection. Such findings are of particular interest, especially at places where intervention studies are difficult to conduct. For example, given the low incidence of new Covid-19 cases in Vietnam as of this writing, an intervention study will require an immense number of participants and/or extended period of follow-up time to determine the vaccine efficacy.

Limitations of these phase 1 and 2 trials are limited ethnic diversity (almost exclusively Vietnamese Kinh people), short follow-up duration, healthy participants only and the lack of convalescent serum data. Without comparison of vaccine responses to convalescent sera from covid-19 patients, it is difficult to compare and contrast the immunogenicity of different vaccines and hence the projected efficacy. This shortcoming will be addressed in phase 3 trial.

In conclusion, Nanocovax is highly safe and immunogenic. Dose strength of 25 mcg is selected for phase 3 to evaluate the vaccine efficacy.

## Data Availability

Data will be provided upon request.

## References

1. COVID Live Update: 190,075,727 Cases and 4,087,371 Deaths from the Coronavirus - Worldometer [Internet]. [cited 2021 Jul 17];Available from: https://www.worldometers.info/coronavirus/

2. Llanes A, Restrepo CM, Caballero Z, Rajeev S, Kennedy MA, Lleonart R. Betacoronavirus Genomes: How Genomic Information has been Used to Deal with Past Outbreaks and the COVID-19 Pandemic. Int J Mol Sci 2020;21(12):E4546.

3. Coronaviridae Study Group of the International Committee on Taxonomy of Viruses. The species Severe acute respiratory syndrome-related coronavirus: classifying 2019-nCoV and naming it SARS-CoV-2. Nat Microbiol 2020;5(4):536–44.

4. Watanabe Y, Allen JD, Wrapp D, McLellan JS, Crispin M. Site-specific glycan analysis of the SARS-CoV-2 spike. Science 2020;369(6501):330–3.

5. Huang Y, Yang C, Xu X, Xu W, Liu S. Structural and functional properties of SARS-CoV-2 spike protein: potential antivirus drug development for COVID-19. Acta Pharmacol Sin 2020;41(9):1141–9.

6. Tran TNM, May B, Ung TT, et al. PRE-CLINICAL IMMUNE RESPONSE AND SAFETY EVALUATION OF THE PROTEIN SUBUNIT VACCINE NANOCOVAX FOR COVID-19. bioRxiv 2021;2021.07.20.453162.

7. Guidance for Industry: Toxicity Grading Scale for Healthy Adult and Adolescent Volunteers Enrolled in Preventive Vaccine Clinical Trials. :10.

8. Division of AIDS (DAIDS) Table for Grading the Severity of Adult and Pediatric Adverse Events. :35.

9. Tan CW, Chia WN, Qin X, et al. A SARS-CoV-2 surrogate virus neutralization test based on antibody-mediated blockage of ACE2–spike protein–protein interaction. Nat Biotechnol 2020;38(9):1073–8.

10. Keech C, Albert G, Cho I, et al. Phase 1–2 Trial of a SARS-CoV-2 Recombinant Spike Protein Nanoparticle Vaccine. N Engl J Med 2020;383(24):2320–32.

11. Chu L, McPhee R, Huang W, et al. A preliminary report of a randomized controlled phase 2 trial of the safety and immunogenicity of mRNA-1273 SARS-CoV-2 vaccine. Vaccine 2021;39(20):2791–9.

12. Sadoff J, Le Gars M, Shukarev G, et al. Interim Results of a Phase 1–2a Trial of Ad26.COV2.S Covid-19 Vaccine. N Engl J Med 2021;384(19):1824–35.

13. Xia S, Zhang Y, Wang Y, et al. Safety and immunogenicity of an inactivated SARS-CoV-2 vaccine, BBIBP-CorV: a randomised, double-blind, placebo-controlled, phase 1/2 trial. The Lancet Infectious Diseases 2021;21(1):39–51.

14. Logunov DY, Dolzhikova IV, Zubkova OV, et al. Safety and immunogenicity of an rAd26 and rAd5 vector-based heterologous prime-boost COVID-19 vaccine in two formulations: two open, non-randomised phase 1/2 studies from Russia. The Lancet 2020;396(10255):887–97.

15. Folegatti PM, Ewer KJ, Aley PK, et al. Safety and immunogenicity of the ChAdOx1 nCoV-19 vaccine against SARS-CoV-2: a preliminary report of a phase 1/2, single-blind, randomised controlled trial. The Lancet 2020;396(10249):467–78.

16. Walsh EE, Frenck RW, Falsey AR, et al. Safety and Immunogenicity of Two RNA-Based Covid-19 Vaccine Candidates. N Engl J Med 2020;383(25):2439–50.

17. Lindblad EB, Schønberg NE. Aluminum adjuvants: preparation, application, dosage, and formulation with antigen. Methods Mol Biol 2010;626:41–58.

18. Arvin AM, Fink K, Schmid MA, et al. A perspective on potential antibody-dependent enhancement of SARS-CoV-2. Nature 2020;584(7821):353–63.

19. Development and Licensure of Vaccines to Prevent COVID-19; Guidance for Industry.:24.

20. Khoury DS, Cromer D, Reynaldi A, et al. Neutralizing antibody levels are highly predictive of immune protection from symptomatic SARS-CoV-2 infection. Nat Med 2021;27(7):1205–11.

