## Supplementary data for "Safety and Immunogenicity of Nanocovax, a SARS-CoV-2 Recombinant Spike Protein Vaccine"

### Supplementary figures and tables

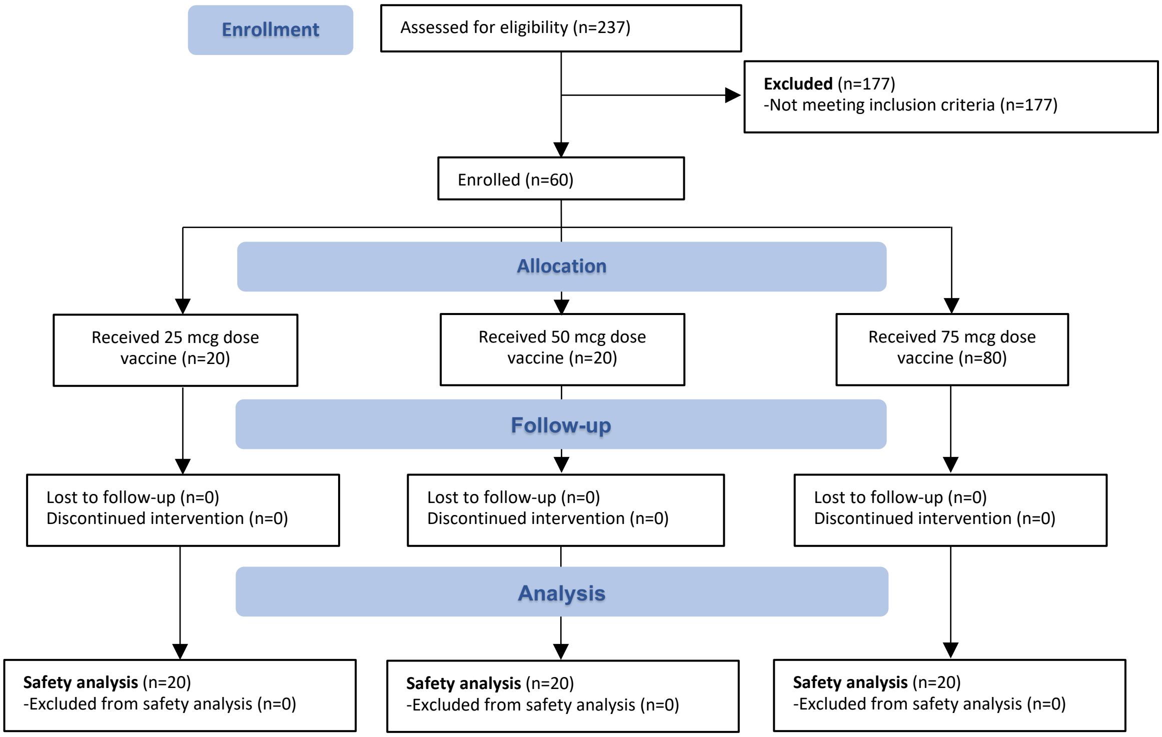


**Figure S1**. Screening and randomization of participants in phase 1.

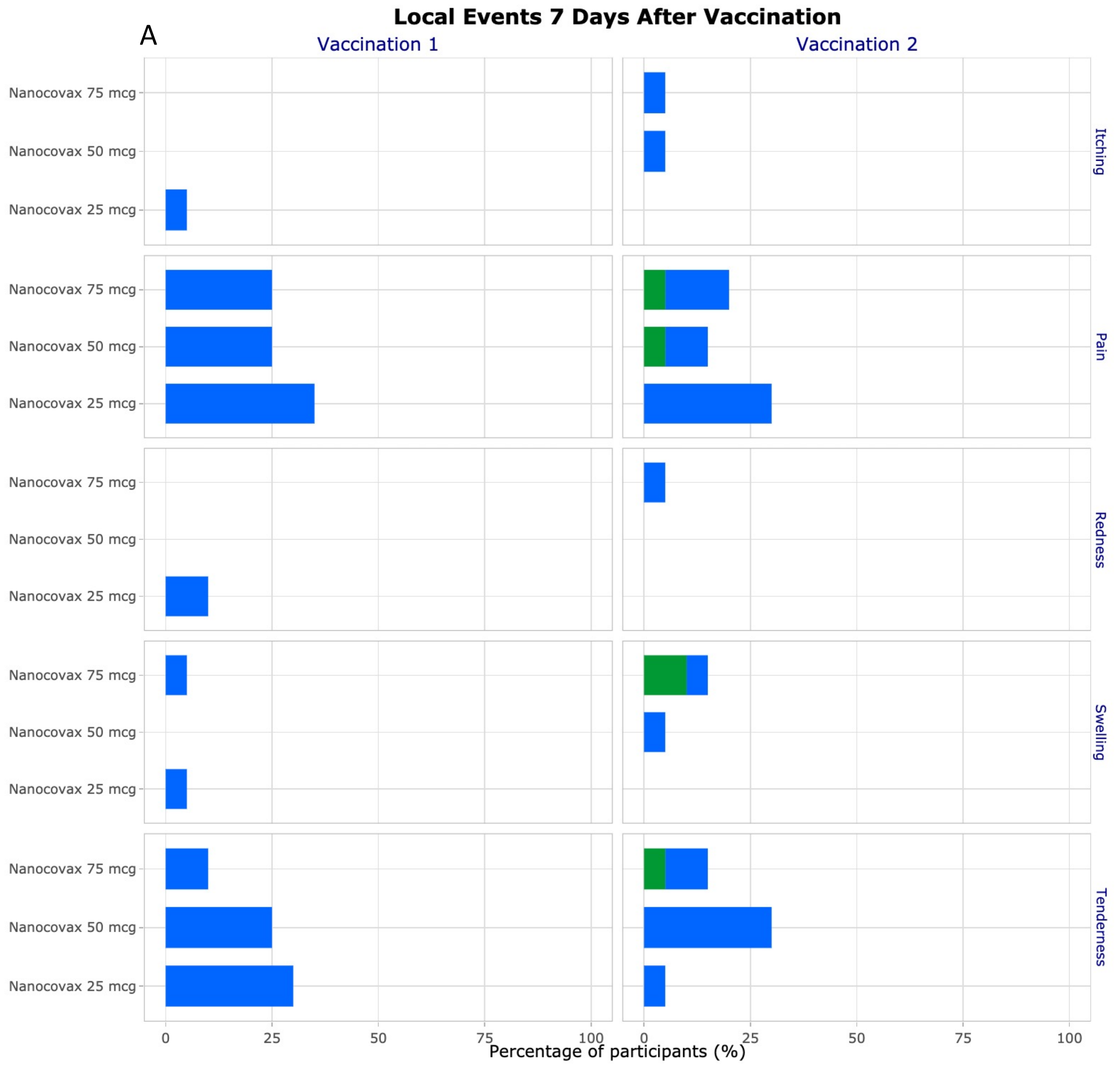
Local Events 7 Days After Vaccination
A
Vaccination 1
Vaccination 2
Nanocovax 75 mcg
Nanocovax 50 mcg
Nanocovax 25 mcg
Itching
Pain
Redness
Swelling
Tenderness
0
25
50
75
100
Percentage of participants (%)

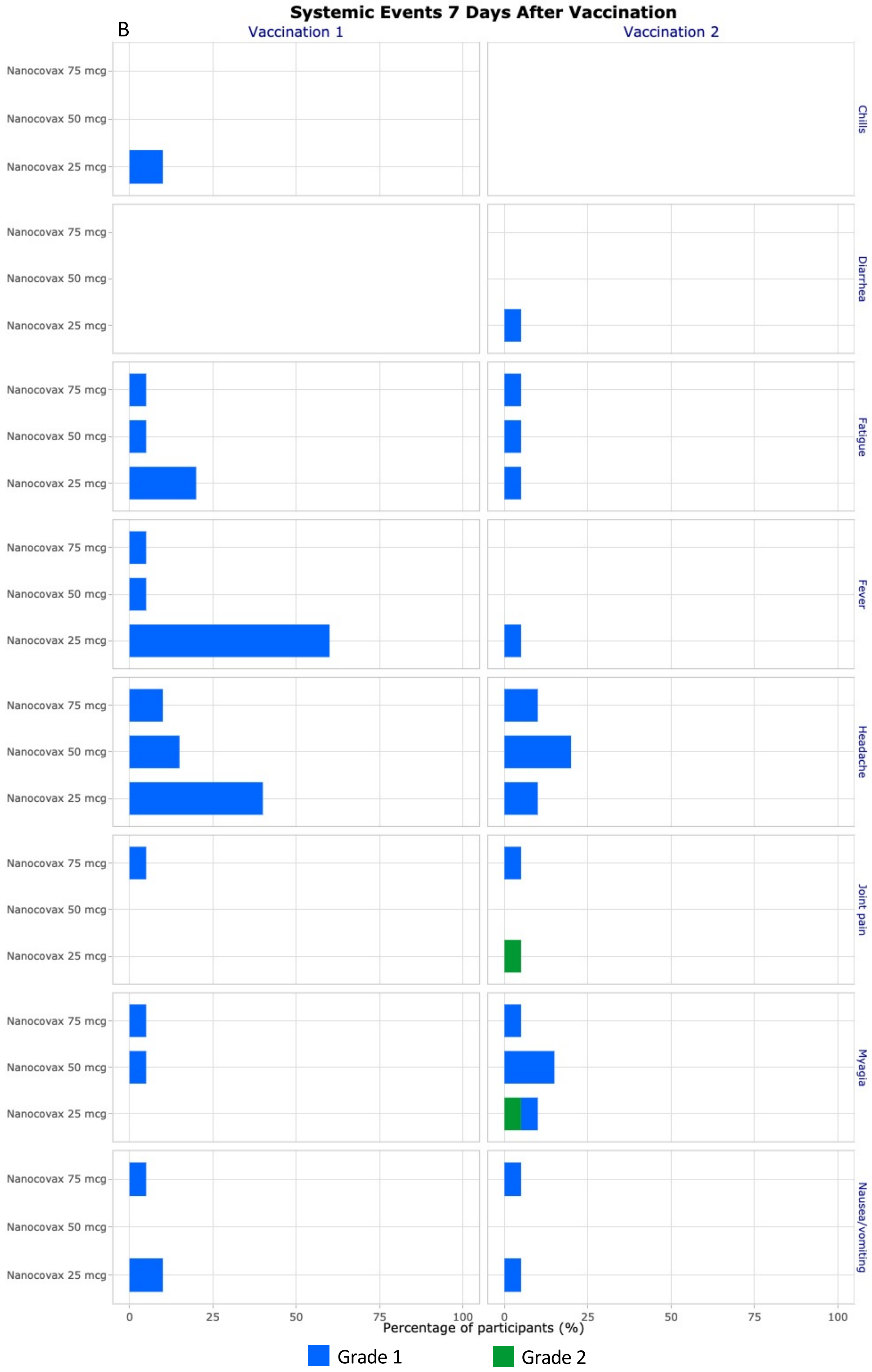


**Figure S2**. Solicited local adverse event (A) and systemic adverse events (B) within 7 days after vaccination in phase 1.

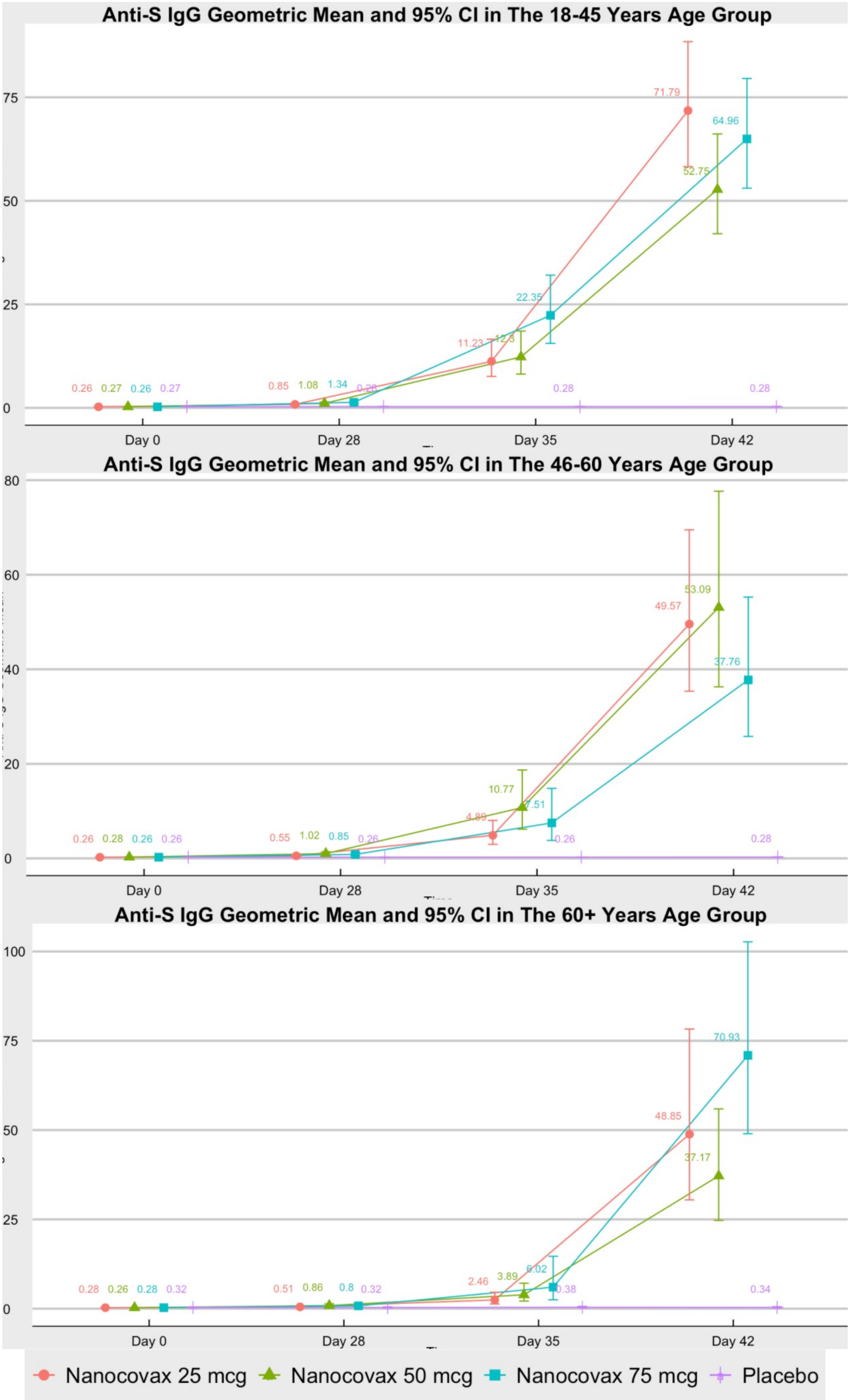


**Figure S3**. Anti-S IgG antibody responses by group age and doses. I bar represents 95% CI.

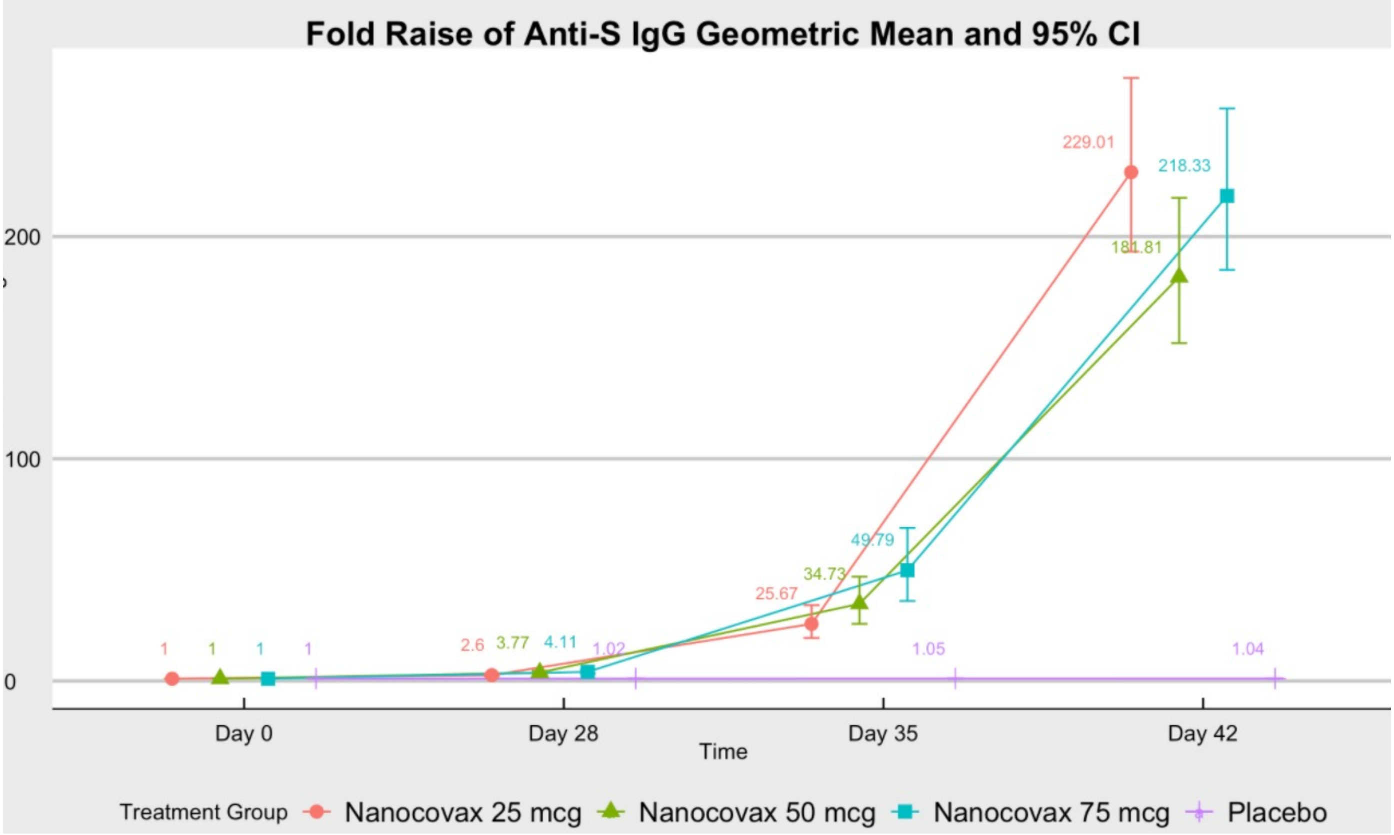


**Figure S4**. Geometric mean fold rise (GMFR) of anti-S IgG at indicated timepoints compared to baseline values (day 0). GMFR is the number of fold increase in GMC of anti-S IgG at a given timepoint compared to its baseline value of the same group on day 0. I bar represents 95% CI.

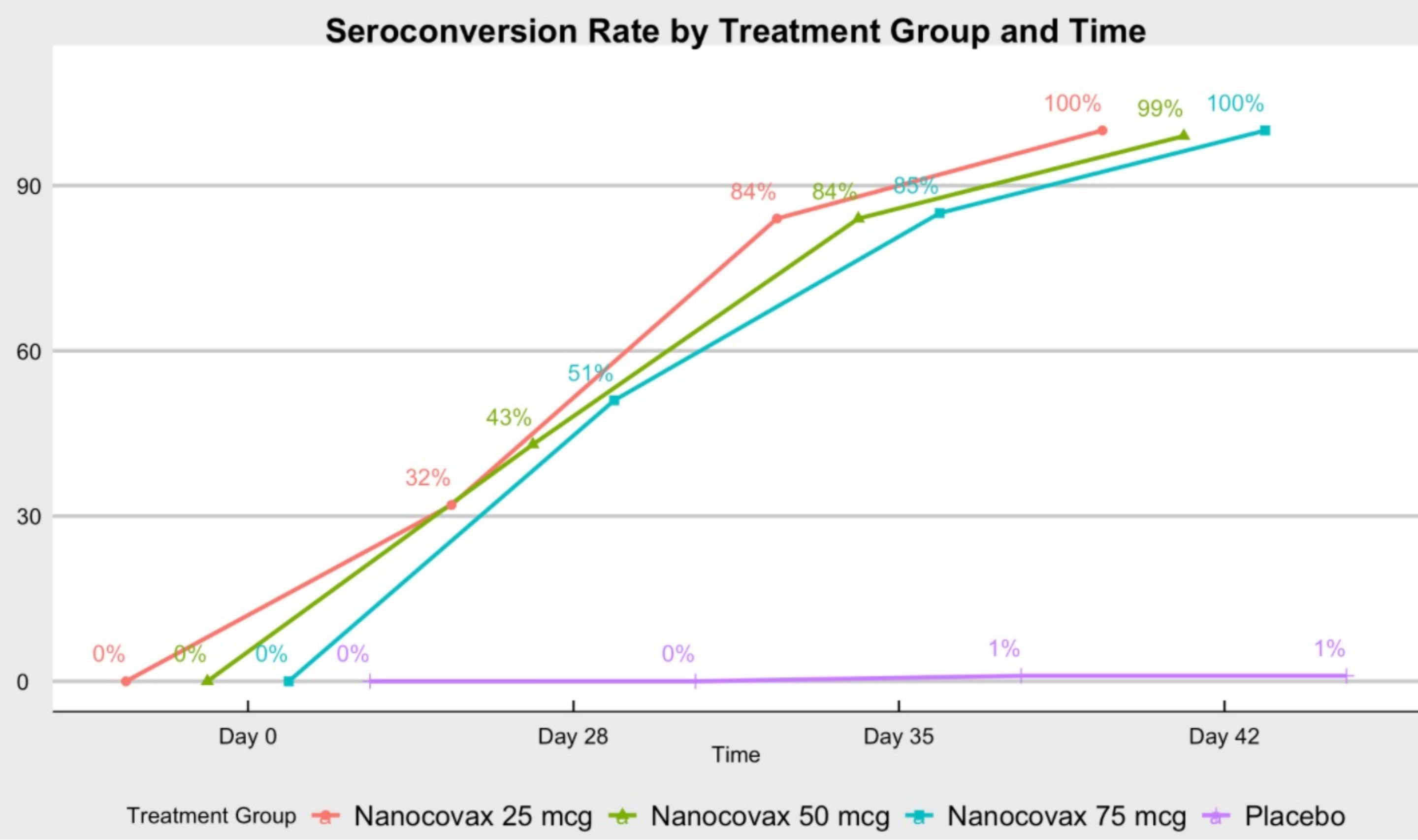


**Figure S5**. Seroconversion rate, defined as GMFR > 4, of vaccine and placebo groups.

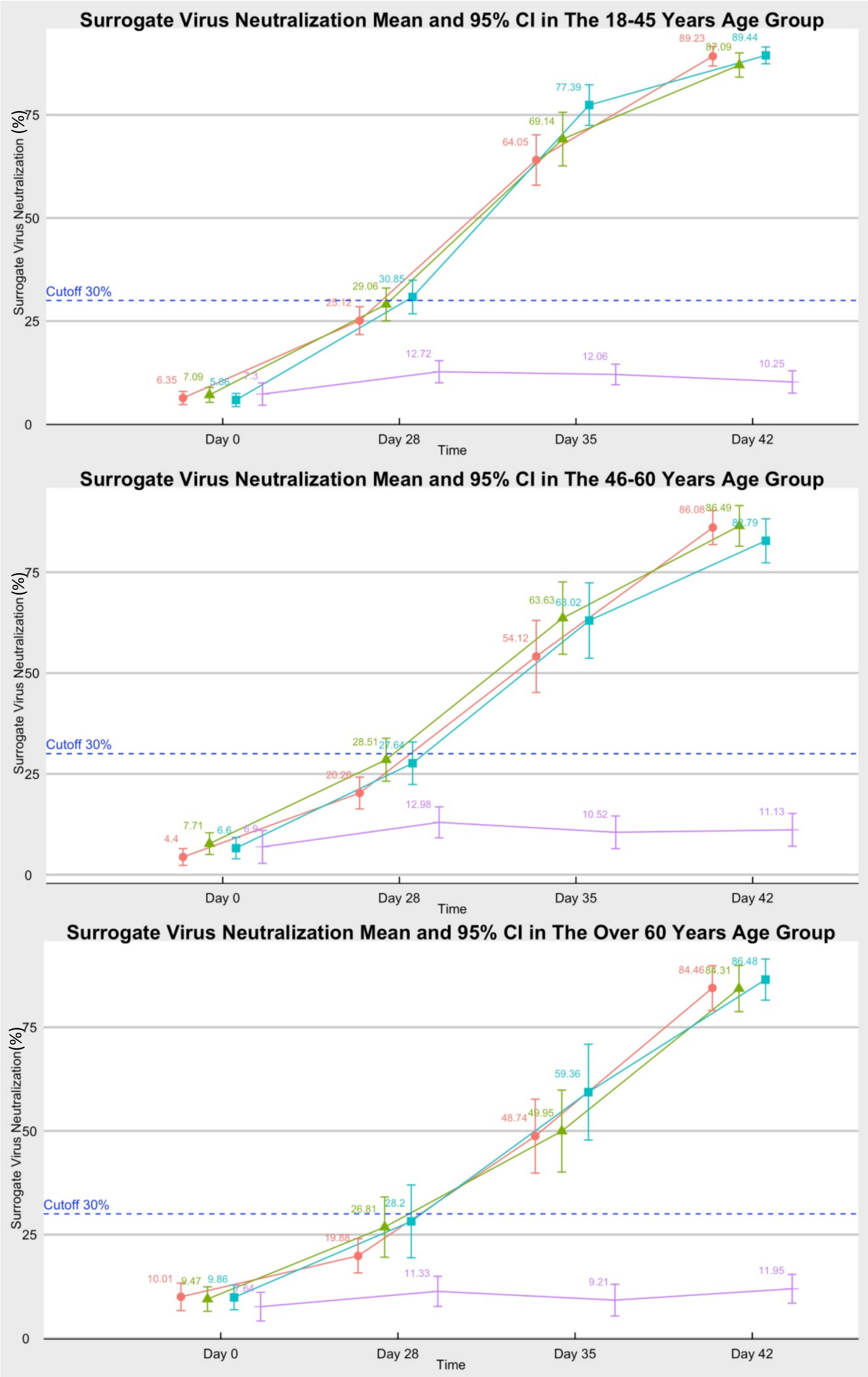


**Figure S6**. Surrogate virus neutralization test results, expressed as inhibition percentage, by group age and doses. I bar represents 95% CI.

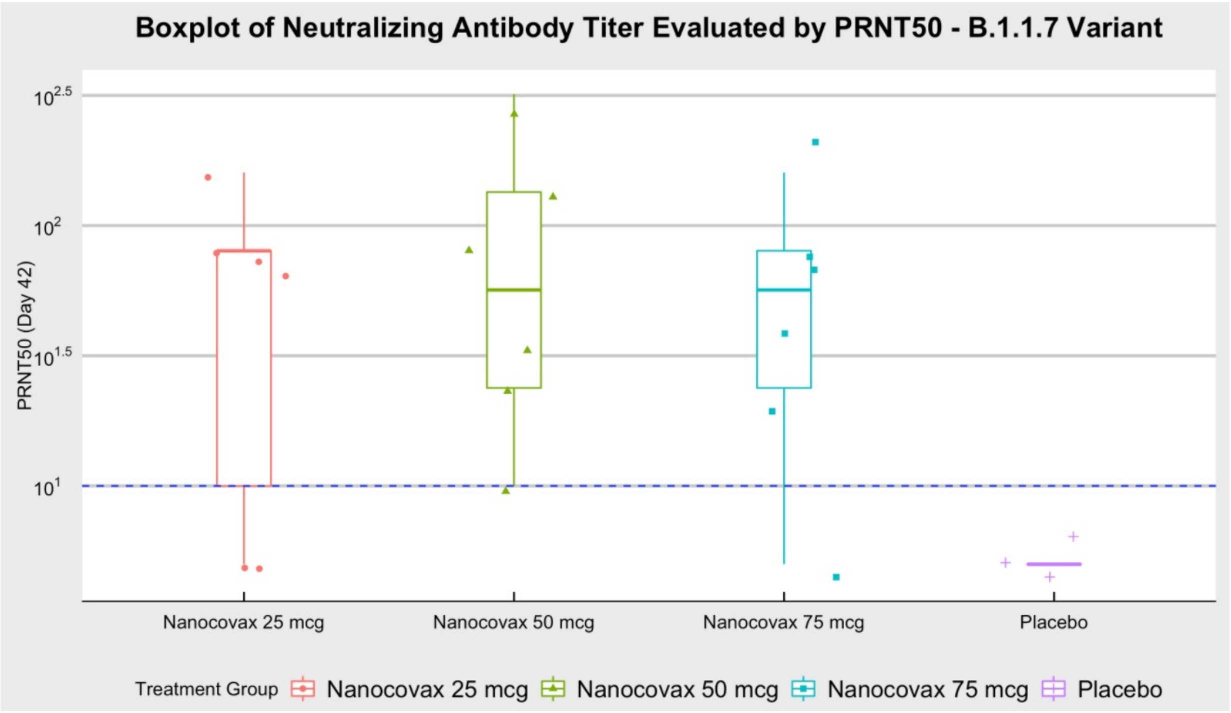


**Figure S7**. $PRNT_{50}$ results of some serum samples collected on day 42 on UK variant (B.1.1.7).

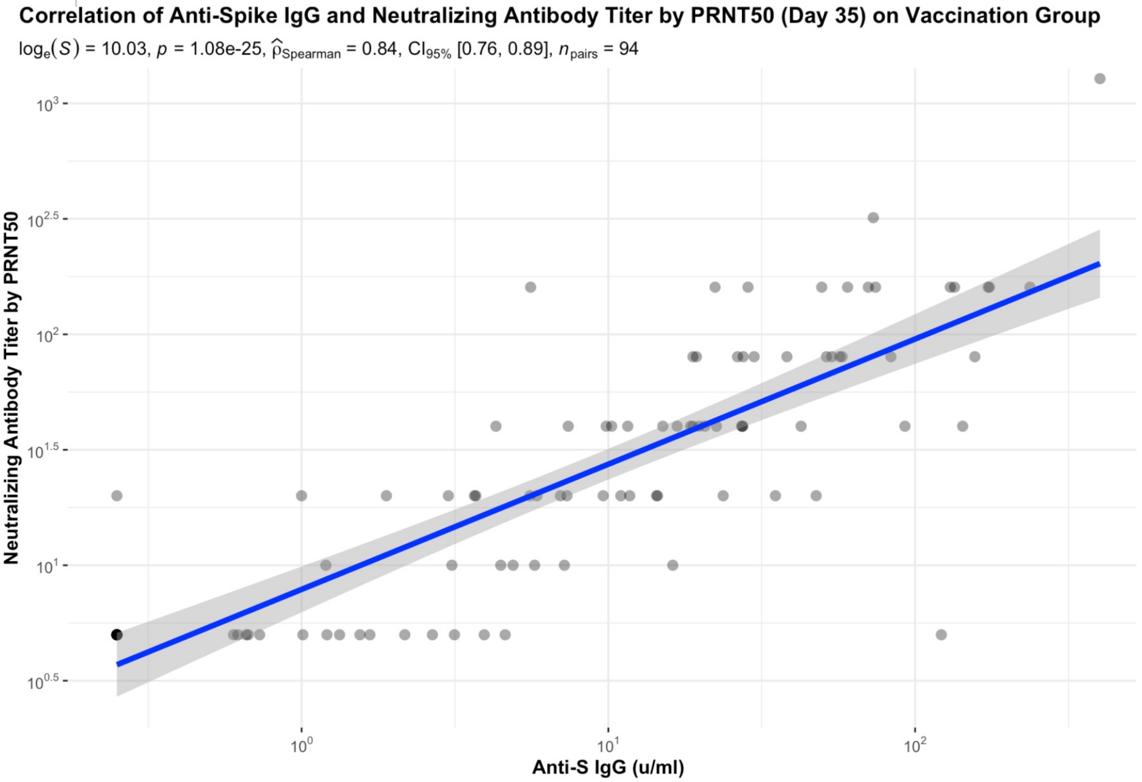


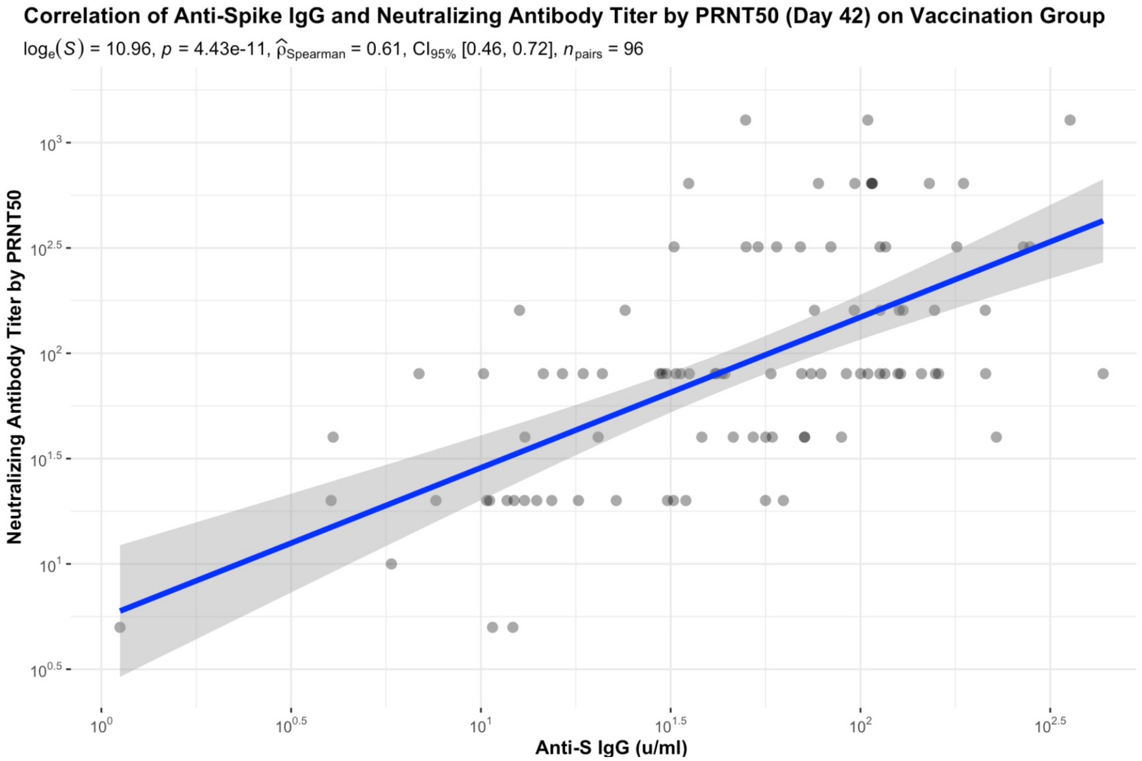


**Figure S8**. Correlation of anti-S IgG and neutralizing antibody responses on day 35 and 42.

**Table S1** Vaccine regimens and key trial timings of phase 1

**A. Vaccine Regimens**

| Vaccine Group | No. of participants | Day 0 | Day 28 |
|---|---|---|---|
| 1.1 | 20 | 25 μg | 25 μg |
| 1.2 | 20 | 50 μg | 50 μg |
| 1.3 | 20 | 75 μg | 75 μg |

**B. Key Trial Timings**

| Procedure | Screening | Day | | | | |
|---|---|---|---|---|---|---|
| | | 0 (1st dose) | 7 | 28 (2nd dose) | 35 | 56 |
| Vaccination | | X | | X | | |
| Follow-up after vaccination | | X | | X | | |
| Vital signs, physical examination | X | X | X | X | X | X |
| Blood sample: safety | X | | X | X | X | X |
| Immunogenicity Assessment | | X | X | X | X | X |
| AEs, concomitant medications | *During the study* | | | | | |

**Table S2**. Demographic characteristics of the participants in the phase 1 trial at enrollment.

| **Characteristics** | **Low dose (25 mcg)** | **Medium dose (50 mcg)** | **High dose (75 mcg)** | **Total** |
|---|---|---|---|---|
| **Group** | **1.1** | **1.2** | **1.3** | |
| Number (n) | 20 | 20 | 20 | 60 |
| **Age (in year)** | | | | |
| Mean ± SD | 23.5 ± 7.1 | 26.6 ± 8.7 | 23.3 ± 3.8 | 24.4 ± 6.9 |
| Range (min - max) | 20 - 43 | 20 - 48 | 21 - 38 | 20 - 48 |
| **Sex – n (%)** | | | | |
| Male | 5 (25%) | 13 (65%) | 5 (25%) | 23 (38%) |
| Female | 15 (75%) | 7 (35%) | 15 (75%) | 37 (62%) |
| **Ethnicity – n (%)** | | | | |
| Kinh | 19 (95%) | 19 (95%) | 20 (100%) | 58 (97%) |
| Other | 1 (5%) | 1 (5%) | 0 | 2 (3%) |
| **Body Mass Index (BMI)*** | | | | |
| Number (n) | 20 | 20 | 20 | 60 |
| Mean ± SD | 21.8 ± 2.1 | 21.5 ± 1.8 | 21.0 ± 1.6 | 21.4 ± 1.9 |
| Range (min - max) | 18.6-26.9 | 18.8-25.5 | 18.5-23.8 | 18.5-26.9 |

**Table S3.** Serious adverse events (SAEs) in phase 2

| No | Site | SAE | Maximum intensity | Outcome |
|---|---|---|---|---|
| 1 | VMMU | Follow up anaphylaxis Grade 1 | - Relation with the investigational product (IP): not confirmed<br>- Severity grade: require medical intervention | Recovered without sequelae |
| 2 | VMMU | Stable angina with stented LCx2 (2017) | - Relation with the IP: not related<br>- Severity grade: hospitalization | Recovered without sequelae |
| 3 | VMMU | Septic fever (Monitor for sepsis) | - Relation with the IP: not related<br>- Severity grade: hospitalization | Recovered without sequelae |
| 4 | Pasteur Institute HCMC | Armpit abscess (R) | - Relation with the IP: not related<br>- Severity grade: hospitalization | Recovered without sequelae |
| 5 | Pasteur Institute HCMC | Injury (broken toe N2+5, wounded N3+4) | - Relation with the IP: not related<br>- Severity grade: hospitalization | Recovered |

#### SUPPLEMENTARY APPENDIX

**1. Vaccination pause rules**

Adverse events meeting any one of the following criteria will result in a hold being placed on subsequent vaccinations pending further review by the safety monitoring committee (SMC):

• Any SAE attributed to vaccine.

• Any toxicity grade 3 (severe) solicited single AE term occurring in ≥ 7 participants across any vaccine dose following vaccination (first and second vaccinations to be assessed separately).

• Toxicity grade 3 (severe) solicited single prespecified laboratory value occurring in ≥ 7 participants across any vaccine dose following injection (first and second vaccinations to be assessed separately). Prespecified laboratory values to be evaluated include creatinine, alanine aminotransferase, aspartate aminotransferase, bilirubin, hemoglobin, complete white blood count, and platelets.

• Any grade 3 (severe) unsolicited single AE preferred term for which the investigator assesses as related which occurs in ≥ 7 participants across any vaccine dose, within 49 days following vaccination (first).

**2. Realtime RT-PCR testing assays**

Real-Time Polymerase Chain Reaction (RT-PCR) for detection of SARS-CoV-2 RNA was performed at the study sites (Vietnam Military Medical Academy) and the Pasteur Institute at Ho Chi Minh city. SARS-CoV-2 RNA was isolated by MagMAX$^{TM}$ Viral/Pathogen nucleic acid isolation kit (#A42352). Primers specifically target RdRP and E genes. The assay can detect SARS-CoV-2 RNA from 1x10^7 copies/reaction to 10 copies/reaction.

**3. Anti-SARS-CoV-2 spike protein serum IgG**

Anti-S IgG in serum samples were evaluated using ADVIA Centaur SARS-CoV-2 IgG (sCOVG) kit (REF# 11207376/11207377) and ADVIA Centaur XP/XPT system by Siemens. All samples were processed according to the manufacturer's procedures with appropriate controls and calibrators by trained laboratory staff. Results of SARS-CoV-2 IgG are given as Index Unit per ml (U/ml), whereby the cut-off for positivity is defined as ≥ 1.0 U/ml. Limit of detection and limit of quantification are both 0.50 U/ml. The range of quantification is 0.5-150.0 U/ml, according to manufacturer's report.

**4. Surrogate virus neutralization assay**

In this assay, the neutralizing activity of anti-S antibody was evaluated by the inhibition of receptor binding domain (RBD) on S protein to its receptor angiotensin-converting enzyme 2

(ACE2) immobilized onto surface of 96 microtiter well plate wells using ELISA based cPass™ SARS-CoV-2 Neutralization Antibody Detection kit (REF # L00847/L00847-5, GenScript). All samples were processed according to the manufacturer's procedures with specific controls. Results are given as inhibition percentage (%) with the cut-off value of 30%. Samples with ≥ 30% inhibition are considered positive for SARS-CoV-2 neutralizing antibody and negative otherwise.

**5. Plaque reduction neutralization test at dilution reducing more than 50% number of plaque ($PRNT_{50}$)**

All serum samples were heat inactivated at $56^0$C for 30 minutes to remove complement and allowed to equilibrate to room temperature prior to processing for neutralization titer. Samples were diluted in duplicate to an initial dilution of 1:5 followed by 1:2 serial dilutions resulting in a 6-dilution series with each well containing 100 µL. All dilutions were performed in DMEM (Gibco, 11965-092), supplemented with 10% (v/v) fetal bovine serum (heat inactivated, Sigma), 1% (v/v) penicillin/streptomycin (Gibco, 15140-122), and 1% (v/v) L-glutamine (2 mM final concentration, Gibco, 2503-149). Dilution plates were then transported into the BSL-3 laboratory and 100 µL of diluted SARS-CoV-2 inoculum was added to each well to result in a multiplicity of infection (MOI) of 0.01 upon transfer to 12-well titer plates. A nontreated, virus-only control and a negative control (mock infection) were included on every plate. SARS-CoV-2 either Wuhan strain or UK variant was prepared at the concentration of 2.5 PFU/microliter and mixed with diluted serum samples at ratio 1:1 (v/v). The sample/virus mixture was then incubated at $37^0$C (5.0% $CO_2$) for 1 hour before transferring to 12-well titer plates with 90% confluent Vero E6 cells. Titer plates were incubated at $37^0$C (5.0% $CO_2$) for 5 days. Plates were then fixed and stained for plaque enumeration. The first sample dilution to show > 50% plaque reduction was reported as the minimum sample dilution required to inhibit (neutralize) >50% of the concentration of SARS-CoV2 tested. Expressed in figures as 50% plaque reduction neutralization (PRNT50).

**6. T cell response by intracellular staining for interferon gamma on CD4+ and CD8+ T cells.**

Peripheral blood mononuclear cells (PBMCs) were isolated using Ficoll-Paque Premium (Cytiva, 17544203) and cryopreserved in fetal bovine serum (Gibco, 10438206) containing 10% (v/v) dimethyl sulfoxide (DMSO) (Sigma, D2650). PBMC were rested 8 hours after thawing. Cells with a viability > 85% proceeded to the following assays. PBMCs were cultured in 96-well U-bottom plates at a density of 1x10^6 cells/well and treated with S1 peptide pools (GensScript, RP30020) at concentration of 2 μg/ml or leukocyte activation cocktail including PMA and Ionomycin (positive control) (BD Biosciences, 550583), or medium only (negative control). After incubation at $37^0$C for 18 hours in the presence of BD GolgiPlug™ (BD Biosciences, 555029), cells were labelled for surface markers CD4, CD8 (BD Biosciences, 340443 and 565310). The intracellular cytokines were detected by antibodies specific for T helper 1 (Th1) cytokine IFNg (BD Biosciences, 557718). The samples were processed using a BD FACSCanto II. Data were analyzed using FACSDiva software by BD.

**Appendix 7: Unsolicited Adverse Event by System Organ Classes, Preferred Term and Severity**

Safety Analysis Set- Placebo Vs. Nanocovax (25 mcg, 50 mcg and 75 mcg)

| **Body System**<br>**Preferred Term**<br>**Severity** | **Nanocovax**<br>**N=480** | **Placebo**<br>**N= 80** | **OVERALL**<br>**N=560** |
|---|---|---|---|
| Any Event | 130 ( 27.1%) | 27 ( 33.8%) | 157 ( 28.0%) |
| Life Threatening | 2 ( 0.4%) | | 2 ( 0.4%) |
| Mild | 103 ( 21.5%) | 21 ( 26.3%) | 124 ( 22.1%) |
| Moderate | 20 ( 4.2%) | 6 ( 7.5%) | 26 ( 4.6%) |
| Severe | 5 ( 1.0%) | | 5 ( 0.9%) |
| Blood and lymphatic system disorders | 7 ( 1.5%) | 2 ( 2.5%) | 9 ( 1.6%) |
| Mild | 4 ( 0.8%) | 1 ( 1.3%) | 5 ( 0.9%) |
| Moderate | 3 ( 0.6%) | 1 ( 1.3%) | 4 ( 0.7%) |
| Anemia | 1 ( 0.2%) | 1 ( 1.3%) | 2 ( 0.4%) |
| Mild | | 1 ( 1.3%) | 1 ( 0.2%) |
| Moderate | 1 ( 0.2%) | | 1 ( 0.2%) |
| Hematuria | 1 ( 0.2%) | | 1 ( 0.2%) |
| Mild | 1 ( 0.2%) | | 1 ( 0.2%) |
| Leukocytes | 6 ( 1.3%) | 2 ( 2.5%) | 8 ( 1.4%) |
| Mild | 4 ( 0.8%) | 1 ( 1.3%) | 5 ( 0.9%) |
| Moderate | 2 ( 0.4%) | 1 ( 1.3%) | 3 ( 0.5%) |
| Cardiac disorders | 6 ( 1.3%) | 1 ( 1.3%) | 7 ( 1.3%) |

*Created by D.D.G with SAS 9.4 on 11MAY2021*
*Program: SAS/folders/myfolders/NANOCOVAXP2/AE/UnSoliAE/AESEV.SAS*

**Appendix 7: Unsolicited Adverse Event by System Organ Classes, Preferred Term and Severity**

Safety Analysis Set- Placebo Vs. Nanocovax (25 mcg, 50 mcg and 75 mcg)

| Body System<br>Preferred Term<br>Severity | Nanocovax<br>N=480 | Placebo<br>N= 80 | OVERALL<br>N=560 |
|---|---|---|---|
| Mild | 5 ( 1.0%) | | 5 ( 0.9%) |
| Moderate | 1 ( 0.2%) | 1 ( 1.3%) | 2 ( 0.4%) |
| Chest pain | 6 ( 1.3%) | | 6 ( 1.1%) |
| Mild | 5 ( 1.0%) | | 5 ( 0.9%) |
| Moderate | 1 ( 0.2%) | | 1 ( 0.2%) |
| Stable angina | | 1 ( 1.3%) | 1 ( 0.2%) |
| Moderate | | 1 ( 1.3%) | 1 ( 0.2%) |
| Tachycardia | 1 ( 0.2%) | | 1 ( 0.2%) |
| Mild | 1 ( 0.2%) | | 1 ( 0.2%) |
| Eye disorders | 4 ( 0.8%) | | 4 ( 0.7%) |
| Mild | 3 ( 0.6%) | | 3 ( 0.5%) |
| Moderate | 1 ( 0.2%) | | 1 ( 0.2%) |
| Conjunctivitis | 1 ( 0.2%) | | 1 ( 0.2%) |
| Moderate | 1 ( 0.2%) | | 1 ( 0.2%) |
| Eyes tearing | 1 ( 0.2%) | | 1 ( 0.2%) |
| Mild | 1 ( 0.2%) | | 1 ( 0.2%) |

*Created by D.D.G with SAS 9.4 on 11MAY2021*
*Program: SAS/folders/myfolders/NANOCOVAXP2/AE/UnSoliAE/AESEV.SAS*

**Appendix 7: Unsolicited Adverse Event by System Organ Classes, Preferred Term and Severity**

Safety Analysis Set- Placebo Vs. Nanocovax (25 mcg, 50 mcg and 75 mcg)

| **Body System**<br>**Preferred Term**<br>**Severity** | **Nanocovax**<br>**N=480** | **Placebo**<br>**N= 80** | **OVERALL**<br>**N=560** |
|---|---|---|---|
| Hordeolum | 1 ( 0.2%) | | 1 ( 0.2%) |
| Mild | 1 ( 0.2%) | | 1 ( 0.2%) |
| Itching- eye area | 1 ( 0.2%) | | 1 ( 0.2%) |
| Mild | 1 ( 0.2%) | | 1 ( 0.2%) |
| Gastrointestinal disorders | 8 ( 1.7%) | 1 ( 1.3%) | 9 ( 1.6%) |
| Mild | 6 ( 1.3%) | 1 ( 1.3%) | 7 ( 1.3%) |
| Moderate | 2 ( 0.4%) | | 2 ( 0.4%) |
| Abdominal pains | 2 ( 0.4%) | | 2 ( 0.4%) |
| Mild | 2 ( 0.4%) | | 2 ( 0.4%) |
| Diarhea | 1 ( 0.2%) | | 1 ( 0.2%) |
| Mild | 1 ( 0.2%) | | 1 ( 0.2%) |
| Dry mouth | 1 ( 0.2%) | | 1 ( 0.2%) |
| Mild | 1 ( 0.2%) | | 1 ( 0.2%) |
| Gastralgia | | 1 ( 1.3%) | 1 ( 0.2%) |
| Mild | | 1 ( 1.3%) | 1 ( 0.2%) |

*Created by D.D.G with SAS 9.4 on 11MAY2021*
*Program: SAS/folders/myfolders/NANOCOVAXP2/AE/UnSoliAE/AESEV.SAS*

**Appendix 7: Unsolicited Adverse Event by System Organ Classes, Preferred Term and Severity**

Safety Analysis Set- Placebo Vs. Nanocovax (25 mcg, 50 mcg and 75 mcg)

| Body System<br>Preferred Term<br>Severity | Nanocovax<br>N=480 | Placebo<br>N= 80 | OVERALL<br>N=560 |
|---|---|---|---|
| Gastritis | 1 ( 0.2%) | | 1 ( 0.2%) |
| Moderate | 1 ( 0.2%) | | 1 ( 0.2%) |
| Left upper quadrant pain | 1 ( 0.2%) | | 1 ( 0.2%) |
| Mild | 1 ( 0.2%) | | 1 ( 0.2%) |
| Toothache | 3 ( 0.6%) | | 3 ( 0.5%) |
| Mild | 2 ( 0.4%) | | 2 ( 0.4%) |
| Moderate | 1 ( 0.2%) | | 1 ( 0.2%) |
| General disorders and administration site conditions | 22 ( 4.6%) | 5 ( 6.3%) | 27 ( 4.8%) |
| Mild | 22 ( 4.6%) | 5 ( 6.3%) | 27 ( 4.8%) |
| Anorexia | 1 ( 0.2%) | | 1 ( 0.2%) |
| Mild | 1 ( 0.2%) | | 1 ( 0.2%) |
| Drowsy | | 1 ( 1.3%) | 1 ( 0.2%) |
| Mild | | 1 ( 1.3%) | 1 ( 0.2%) |
| Dyspepsia flatulence | 2 ( 0.4%) | | 2 ( 0.4%) |
| Mild | 2 ( 0.4%) | | 2 ( 0.4%) |

*Created by D.D.G with SAS 9.4 on 11MAY2021*
*Program: SAS/folders/myfolders/NANOCOVAXP2/AE/UnSoliAE/AESEV.SAS*

**Appendix 7: Unsolicited Adverse Event by System Organ Classes, Preferred Term and Severity**

Safety Analysis Set- Placebo Vs. Nanocovax (25 mcg, 50 mcg and 75 mcg)

| Body System<br>Preferred Term<br>Severity | Nanocovax<br>N=480 | Placebo<br>N= 80 | OVERALL<br>N=560 |
|---|---|---|---|
| Fatigue | 2 ( 0.4%) | 1 ( 1.3%) | 3 ( 0.5%) |
| Mild | 2 ( 0.4%) | 1 ( 1.3%) | 3 ( 0.5%) |
| Fever | 5 ( 1.0%) | 1 ( 1.3%) | 6 ( 1.1%) |
| Mild | 5 ( 1.0%) | 1 ( 1.3%) | 6 ( 1.1%) |
| Headache | 1 ( 0.2%) | | 1 ( 0.2%) |
| Mild | 1 ( 0.2%) | | 1 ( 0.2%) |
| Hypothermia | 10 ( 2.1%) | 2 ( 2.5%) | 12 ( 2.1%) |
| Mild | 10 ( 2.1%) | 2 ( 2.5%) | 12 ( 2.1%) |
| Insomnia | 2 ( 0.4%) | | 2 ( 0.4%) |
| Mild | 2 ( 0.4%) | | 2 ( 0.4%) |
| Loss of taste | 1 ( 0.2%) | | 1 ( 0.2%) |
| Mild | 1 ( 0.2%) | | 1 ( 0.2%) |
| Tenderness | 1 ( 0.2%) | | 1 ( 0.2%) |
| Mild | 1 ( 0.2%) | | 1 ( 0.2%) |

*Created by D.D.G with SAS 9.4 on 11MAY2021*
*Program: SAS/folders/myfolders/NANOCOVAXP2/AE/UnSoliAE/AESEV.SAS*

**Appendix 7: Unsolicited Adverse Event by System Organ Classes, Preferred Term and Severity**

Safety Analysis Set- Placebo Vs. Nanocovax (25 mcg, 50 mcg and 75 mcg)

| **Body System<br>Preferred Term<br>Severity** | **Nanocovax<br>N=480** | **Placebo<br>N= 80** | **OVERALL<br>N=560** |
|---|---|---|---|
| Immune system disorders | 3 ( 0.6%) | | 3 ( 0.5%) |
| Mild | 3 ( 0.6%) | | 3 ( 0.5%) |
| Allergic reaction | 2 ( 0.4%) | | 2 ( 0.4%) |
| Mild | 2 ( 0.4%) | | 2 ( 0.4%) |
| Anaphylaxis- Grade I | 1 ( 0.2%) | | 1 ( 0.2%) |
| Mild | 1 ( 0.2%) | | 1 ( 0.2%) |
| Infections and infestations | 3 ( 0.6%) | 1 ( 1.3%) | 4 ( 0.7%) |
| Life Threatening | 1 ( 0.2%) | | 1 ( 0.2%) |
| Mild | 1 ( 0.2%) | 1 ( 1.3%) | 2 ( 0.4%) |
| Severe | 1 ( 0.2%) | | 1 ( 0.2%) |
| Atrioventricular node abscess | 1 ( 0.2%) | | 1 ( 0.2%) |
| Life Threatening | 1 ( 0.2%) | | 1 ( 0.2%) |
| Flu | 1 ( 0.2%) | 1 ( 1.3%) | 2 ( 0.4%) |
| Mild | 1 ( 0.2%) | 1 ( 1.3%) | 2 ( 0.4%) |
| Sepsis | 1 ( 0.2%) | | 1 ( 0.2%) |
| Severe | 1 ( 0.2%) | | 1 ( 0.2%) |

*Created by D.D.G with SAS 9.4 on 11MAY2021*
*Program: SAS/folders/myfolders/NANOCOVAXP2/AE/UnSoliAE/AESEV.SAS*

**Appendix 7: Unsolicited Adverse Event by System Organ Classes, Preferred Term and Severity**

Safety Analysis Set- Placebo Vs. Nanocovax (25 mcg, 50 mcg and 75 mcg)

| **Body System**<br>**Preferred Term**<br>**Severity** | **Nanocovax**<br>**N=480** | **Placebo**<br>**N= 80** | **OVERALL**<br>**N=560** |
|---|---|---|---|
| Injury, poisoning and procedural complications | 1 ( 0.2%) | | 1 ( 0.2%) |
| Moderate | 1 ( 0.2%) | | 1 ( 0.2%) |
| Broken Bone, Right Heel | 1 ( 0.2%) | | 1 ( 0.2%) |
| Moderate | 1 ( 0.2%) | | 1 ( 0.2%) |
| Investigations | 6 ( 1.3%) | 3 ( 3.8%) | 9 ( 1.6%) |
| Mild | 4 ( 0.8%) | 1 ( 1.3%) | 5 ( 0.9%) |
| Moderate | 2 ( 0.4%) | 2 ( 2.5%) | 4 ( 0.7%) |
| Elevated White Blood Cells | 5 ( 1.0%) | 1 ( 1.3%) | 6 ( 1.1%) |
| Mild | 4 ( 0.8%) | 1 ( 1.3%) | 5 ( 0.9%) |
| Moderate | 1 ( 0.2%) | | 1 ( 0.2%) |
| Elevated liver function tests | | 2 ( 2.5%) | 2 ( 0.4%) |
| Moderate | | 2 ( 2.5%) | 2 ( 0.4%) |
| WBC formula shifted | 1 ( 0.2%) | | 1 ( 0.2%) |
| Moderate | 1 ( 0.2%) | | 1 ( 0.2%) |
| Metabolism and nutrition disorders | 11 ( 2.3%) | 2 ( 2.5%) | 13 ( 2.3%) |

*Created by D.D.G with SAS 9.4 on 11MAY2021*
*Program: SAS/folders/myfolders/NANOCOVAXP2/AE/UnSoliAE/AESEV.SAS*

**Appendix 7: Unsolicited Adverse Event by System Organ Classes, Preferred Term and Severity**

Safety Analysis Set- Placebo Vs. Nanocovax (25 mcg, 50 mcg and 75 mcg)

| Body System<br>Preferred Term<br>Severity | Nanocovax<br>N=480 | Placebo<br>N= 80 | OVERALL<br>N=560 |
|---|---|---|---|
| Mild | 11 ( 2.3%) | 2 ( 2.5%) | 13 ( 2.3%) |
| Hyperglycemia | 11 ( 2.3%) | 2 ( 2.5%) | 13 ( 2.3%) |
| Mild | 10 ( 2.1%) | 2 ( 2.5%) | 12 ( 2.1%) |
| Moderate | 1 ( 0.2%) | | 1 ( 0.2%) |
| Urinary glucose | 5 ( 1.0%) | 1 ( 1.3%) | 6 ( 1.1%) |
| Mild | 5 ( 1.0%) | 1 ( 1.3%) | 6 ( 1.1%) |
| Musculoskeletal and connective tissue disorders | 21 ( 4.4%) | 11 ( 13.8%) | 32 ( 5.7%) |
| Life Threatening | 1 ( 0.2%) | | 1 ( 0.2%) |
| Mild | 14 ( 2.9%) | 9 ( 11.3%) | 23 ( 4.1%) |
| Moderate | 4 ( 0.8%) | 2 ( 2.5%) | 6 ( 1.1%) |
| Severe | 2 ( 0.4%) | | 2 ( 0.4%) |
| 2-sided breast pain | 1 ( 0.2%) | | 1 ( 0.2%) |
| Mild | 1 ( 0.2%) | | 1 ( 0.2%) |
| Arthralgia | 6 ( 1.3%) | 3 ( 3.8%) | 9 ( 1.6%) |
| Mild | 6 ( 1.3%) | 2 ( 2.5%) | 8 ( 1.4%) |
| Moderate | | 1 ( 1.3%) | 1 ( 0.2%) |

*Created by D.D.G with SAS 9.4 on 11MAY2021*
*Program: SAS/folders/myfolders/NANOCOVAXP2/AE/UnSoliAE/AESEV.SAS*

**Appendix 7: Unsolicited Adverse Event by System Organ Classes, Preferred Term and Severity**

Safety Analysis Set- Placebo Vs. Nanocovax (25 mcg, 50 mcg and 75 mcg)

| Body System<br>Preferred Term<br>Severity | Nanocovax<br>N=480 | Placebo<br>N= 80 | OVERALL<br>N=560 |
|---|---|---|---|
| Arthritis | | 2 ( 2.5%) | 2 ( 0.4%) |
| Mild | | 1 ( 1.3%) | 1 ( 0.2%) |
| Moderate | | 1 ( 1.3%) | 1 ( 0.2%) |
| Back pain | 2 ( 0.4%) | 1 ( 1.3%) | 3 ( 0.5%) |
| Mild | | 1 ( 1.3%) | 1 ( 0.2%) |
| Moderate | 1 ( 0.2%) | | 1 ( 0.2%) |
| Severe | 1 ( 0.2%) | | 1 ( 0.2%) |
| Cervical scapulohumeral syndrome | 1 ( 0.2%) | | 1 ( 0.2%) |
| Moderate | 1 ( 0.2%) | | 1 ( 0.2%) |
| Degenerative Spine | 2 ( 0.4%) | | 2 ( 0.4%) |
| Mild | 1 ( 0.2%) | | 1 ( 0.2%) |
| Severe | 1 ( 0.2%) | | 1 ( 0.2%) |
| Disc herniation | 1 ( 0.2%) | | 1 ( 0.2%) |
| Moderate | 1 ( 0.2%) | | 1 ( 0.2%) |
| Hand pain | 2 ( 0.4%) | 1 ( 1.3%) | 3 ( 0.5%) |
| Mild | 2 ( 0.4%) | 1 ( 1.3%) | 3 ( 0.5%) |

**Appendix 7: Unsolicited Adverse Event by System Organ Classes, Preferred Term and Severity**

Safety Analysis Set- Placebo Vs. Nanocovax (25 mcg, 50 mcg and 75 mcg)

| **Body System**<br>**Preferred Term**<br>**Severity** | **Nanocovax**<br>**N=480** | **Placebo**<br>**N= 80** | **OVERALL**<br>**N=560** |
|---|---|---|---|
| Knee pain | 1 ( 0.2%) | 1 ( 1.3%) | 2 ( 0.4%) |
| Mild | 1 ( 0.2%) | 1 ( 1.3%) | 2 ( 0.4%) |
| Myalgia | 5 ( 1.0%) | | 5 ( 0.9%) |
| Mild | 5 ( 1.0%) | | 5 ( 0.9%) |
| Neck shoulder pain | 1 ( 0.2%) | 3 ( 3.8%) | 4 ( 0.7%) |
| Life Threatening | 1 ( 0.2%) | | 1 ( 0.2%) |
| Mild | | 3 ( 3.8%) | 3 ( 0.5%) |
| Osteoarthritis | 1 ( 0.2%) | | 1 ( 0.2%) |
| Mild | 1 ( 0.2%) | | 1 ( 0.2%) |
| Rheumatoid arthritis | 1 ( 0.2%) | 1 ( 1.3%) | 2 ( 0.4%) |
| Moderate | 1 ( 0.2%) | 1 ( 1.3%) | 2 ( 0.4%) |
| Sore feet | | 1 ( 1.3%) | 1 ( 0.2%) |
| Mild | | 1 ( 1.3%) | 1 ( 0.2%) |
| Spinal muscle pain | 1 ( 0.2%) | | 1 ( 0.2%) |
| Mild | 1 ( 0.2%) | | 1 ( 0.2%) |

**Appendix 7: Unsolicited Adverse Event by System Organ Classes, Preferred Term and Severity**

Safety Analysis Set- Placebo Vs. Nanocovax (25 mcg, 50 mcg and 75 mcg)

| **Body System<br>Preferred Term<br>Severity** | **Nanocovax<br>N=480** | **Placebo<br>N= 80** | **OVERALL<br>N=560** |
|---|---|---|---|
| Spinal pain | 2 ( 0.4%) | | 2 ( 0.4%) |
| Mild | 2 ( 0.4%) | | 2 ( 0.4%) |
| Nervous system disorders | 7 ( 1.5%) | 3 ( 3.8%) | 10 ( 1.8%) |
| Life Threatening | 1 ( 0.2%) | | 1 ( 0.2%) |
| Mild | 5 ( 1.0%) | 3 ( 3.8%) | 8 ( 1.4%) |
| Moderate | 1 ( 0.2%) | | 1 ( 0.2%) |
| Dizziness | 4 ( 0.8%) | 2 ( 2.5%) | 6 ( 1.1%) |
| Life Threatening | 1 ( 0.2%) | | 1 ( 0.2%) |
| Mild | 1 ( 0.2%) | 2 ( 2.5%) | 3 ( 0.5%) |
| Moderate | 2 ( 0.4%) | | 2 ( 0.4%) |
| Headache | 5 ( 1.0%) | | 5 ( 0.9%) |
| Mild | 5 ( 1.0%) | | 5 ( 0.9%) |
| Vestibular disorders | | 1 ( 1.3%) | 1 ( 0.2%) |
| Mild | | 1 ( 1.3%) | 1 ( 0.2%) |
| Renal and urinary disorders | 12 ( 2.5%) | 2 ( 2.5%) | 14 ( 2.5%) |
| Mild | 5 ( 1.0%) | 1 ( 1.3%) | 6 ( 1.1%) |
| Moderate | 7 ( 1.5%) | 1 ( 1.3%) | 8 ( 1.4%) |

*Created by D.D.G with SAS 9.4 on 11MAY2021*
*Program: SAS/folders/myfolders/NANOCOVAXP2/AE/UnSoliAE/AESEV.SAS*

**Appendix 7: Unsolicited Adverse Event by System Organ Classes, Preferred Term and Severity**

Safety Analysis Set- Placebo Vs. Nanocovax (25 mcg, 50 mcg and 75 mcg)

| Body System<br>Preferred Term<br>Severity | Nanocovax<br>N=480 | Placebo<br>N= 80 | OVERALL<br>N=560 |
|---|---|---|---|
| Cystitis | 1 ( 0.2%) | | 1 ( 0.2%) |
| Moderate | 1 ( 0.2%) | | 1 ( 0.2%) |
| Hematuria | 7 ( 1.5%) | 2 ( 2.5%) | 9 ( 1.6%) |
| Mild | 2 ( 0.4%) | 1 ( 1.3%) | 3 ( 0.5%) |
| Moderate | 5 ( 1.0%) | 1 ( 1.3%) | 6 ( 1.1%) |
| Painful urination | 1 ( 0.2%) | | 1 ( 0.2%) |
| Moderate | 1 ( 0.2%) | | 1 ( 0.2%) |
| Urinary tract infection | 3 ( 0.6%) | | 3 ( 0.5%) |
| Mild | 3 ( 0.6%) | | 3 ( 0.5%) |
| Reproductive system and breast disorders | 2 ( 0.4%) | 2 ( 2.5%) | 4 ( 0.7%) |
| Mild | 2 ( 0.4%) | 1 ( 1.3%) | 3 ( 0.5%) |
| Moderate | | 1 ( 1.3%) | 1 ( 0.2%) |
| Menstruation irregular | 2 ( 0.4%) | 2 ( 2.5%) | 4 ( 0.7%) |
| Mild | 2 ( 0.4%) | 1 ( 1.3%) | 3 ( 0.5%) |
| Moderate | | 1 ( 1.3%) | 1 ( 0.2%) |

*Created by D.D.G with SAS 9.4 on 11MAY2021*
*Program: SAS/folders/myfolders/NANOCOVAXP2/AE/UnSoliAE/AESEV.SAS*

**Appendix 7: Unsolicited Adverse Event by System Organ Classes, Preferred Term and Severity**

Safety Analysis Set- Placebo Vs. Nanocovax (25 mcg, 50 mcg and 75 mcg)

| Body System<br>Preferred Term<br>Severity | Nanocovax<br>N=480 | Placebo<br>N= 80 | OVERALL<br>N=560 |
|---|---|---|---|
| Respiratory, thoracic and mediastinal disorders | 43 ( 9.0%) | 6 ( 7.5%) | 49 ( 8.8%) |
| Mild | 37 ( 7.7%) | 6 ( 7.5%) | 43 ( 7.7%) |
| Moderate | 5 ( 1.0%) | | 5 ( 0.9%) |
| Severe | 1 ( 0.2%) | | 1 ( 0.2%) |
| Acute sinusitis | 1 ( 0.2%) | | 1 ( 0.2%) |
| Moderate | 1 ( 0.2%) | | 1 ( 0.2%) |
| Allergic Nasopharyngitis | 1 ( 0.2%) | | 1 ( 0.2%) |
| Moderate | 1 ( 0.2%) | | 1 ( 0.2%) |
| Allergic rhinitis | 1 ( 0.2%) | | 1 ( 0.2%) |
| Mild | 1 ( 0.2%) | | 1 ( 0.2%) |
| Amidan swelling | 1 ( 0.2%) | | 1 ( 0.2%) |
| Moderate | 1 ( 0.2%) | | 1 ( 0.2%) |
| Cough | 9 ( 1.9%) | 2 ( 2.5%) | 11 ( 2.0%) |
| Mild | 9 ( 1.9%) | 2 ( 2.5%) | 11 ( 2.0%) |
| Dyspnea | 2 ( 0.4%) | | 2 ( 0.4%) |
| Mild | 1 ( 0.2%) | | 1 ( 0.2%) |

*Created by D.D.G with SAS 9.4 on 11MAY2021*
*Program: SAS/folders/myfolders/NANOCOVAXP2/AE/UnSoliAE/AESEV.SAS*

**Appendix 7: Unsolicited Adverse Event by System Organ Classes, Preferred Term and Severity**

Safety Analysis Set- Placebo Vs. Nanocovax (25 mcg, 50 mcg and 75 mcg)

| **Body System<br>Preferred Term<br>Severity** | **Nanocovax<br>N=480** | **Placebo<br>N= 80** | **OVERALL<br>N=560** |
|---|---|---|---|
| Moderate | 1 ( 0.2%) | | 1 ( 0.2%) |
| Hoarseness | 1 ( 0.2%) | | 1 ( 0.2%) |
| Mild | 1 ( 0.2%) | | 1 ( 0.2%) |
| Nasal Discharge | 6 ( 1.3%) | 2 ( 2.5%) | 8 ( 1.4%) |
| Mild | 6 ( 1.3%) | 2 ( 2.5%) | 8 ( 1.4%) |
| Nasal pain | | 1 ( 1.3%) | 1 ( 0.2%) |
| Mild | | 1 ( 1.3%) | 1 ( 0.2%) |
| Sinusitis | 1 ( 0.2%) | | 1 ( 0.2%) |
| Mild | 1 ( 0.2%) | | 1 ( 0.2%) |
| Sore Throat | 23 ( 4.8%) | 4 ( 5.0%) | 27 ( 4.8%) |
| Mild | 20 ( 4.2%) | 4 ( 5.0%) | 24 ( 4.3%) |
| Moderate | 2 ( 0.4%) | | 2 ( 0.4%) |
| Severe | 1 ( 0.2%) | | 1 ( 0.2%) |
| Throat sensation disorders | 2 ( 0.4%) | | 2 ( 0.4%) |
| Mild | 2 ( 0.4%) | | 2 ( 0.4%) |

*Created by D.D.G with SAS 9.4 on 11MAY2021*
*Program: SAS/folders/myfolders/NANOCOVAXP2/AE/UnSoliAE/AESEV.SAS*

**Appendix 7: Unsolicited Adverse Event by System Organ Classes, Preferred Term and Severity**

Safety Analysis Set- Placebo Vs. Nanocovax (25 mcg, 50 mcg and 75 mcg)

| Body System<br>Preferred Term<br>Severity | Nanocovax<br>N=480 | Placebo<br>N= 80 | OVERALL<br>N=560 |
|---|---|---|---|
| Upper respiratory tract infection | 1 ( 0.2%) | | 1 ( 0.2%) |
| Mild | 1 ( 0.2%) | | 1 ( 0.2%) |
| Skin and subcutaneous tissue disorders | 6 ( 1.3%) | 2 ( 2.5%) | 8 ( 1.4%) |
| Mild | 5 ( 1.0%) | 2 ( 2.5%) | 7 ( 1.3%) |
| Moderate | 1 ( 0.2%) | | 1 ( 0.2%) |
| Burn | 1 ( 0.2%) | | 1 ( 0.2%) |
| Moderate | 1 ( 0.2%) | | 1 ( 0.2%) |
| Itching- head skin | | 1 ( 1.3%) | 1 ( 0.2%) |
| Mild | | 1 ( 1.3%) | 1 ( 0.2%) |
| Itchy rash | 3 ( 0.6%) | 1 ( 1.3%) | 4 ( 0.7%) |
| Mild | 3 ( 0.6%) | 1 ( 1.3%) | 4 ( 0.7%) |
| Lumps at the injection site | 1 ( 0.2%) | | 1 ( 0.2%) |
| Mild | 1 ( 0.2%) | | 1 ( 0.2%) |
| Skin infection | 1 ( 0.2%) | | 1 ( 0.2%) |
| Mild | 1 ( 0.2%) | | 1 ( 0.2%) |

*Created by D.D.G with SAS 9.4 on 11MAY2021*
*Program: SAS/folders/myfolders/NANOCOVAXP2/AE/UnSoliAE/AESEV.SAS*

**Appendix 7: Unsolicited Adverse Event by System Organ Classes, Preferred Term and Severity**

Safety Analysis Set- Placebo Vs. Nanocovax (25 mcg, 50 mcg and 75 mcg)

| **Body System<br>Preferred Term<br>Severity** | **Nanocovax<br>N=480** | **Placebo<br>N= 80** | **OVERALL<br>N=560** |
|---|---|---|---|
| Vascular disorders | 6 ( 1.3%) | 2 ( 2.5%) | 8 ( 1.4%) |
| Mild | 4 ( 0.8%) | 1 ( 1.3%) | 5 ( 0.9%) |
| Moderate | 1 ( 0.2%) | 1 ( 1.3%) | 2 ( 0.4%) |
| Severe | 1 ( 0.2%) | | 1 ( 0.2%) |
| Hypertension | 6 ( 1.3%) | 1 ( 1.3%) | 7 ( 1.3%) |
| Mild | 4 ( 0.8%) | | 4 ( 0.7%) |
| Moderate | 1 ( 0.2%) | 1 ( 1.3%) | 2 ( 0.4%) |
| Severe | 1 ( 0.2%) | | 1 ( 0.2%) |
| Hypotension | | 1 ( 1.3%) | 1 ( 0.2%) |
| Mild | | 1 ( 1.3%) | 1 ( 0.2%) |

*Created by D.D.G with SAS 9.4 on 11MAY2021*
*Program: SAS/folders/myfolders/NANOCOVAXP2/AE/UnSoliAE/AESEV.SAS*

**Appendix 7: Unsolicited Adverse Events by System Organ Classes, Preferred Term and Relationship to Study Drug**

Safety Analysis Set- Placebo Vs. Nanocovax (25mcg/50mcg/75mcg

| Body System<br>Preferred Term<br>Relationship | Nanocovax<br>N=480 | Placebo<br>N= 80 | OVERALL<br>N=560 |
|---|---|---|---|
| Any Event | 130 ( 27.1%) | 27 ( 33.8%) | 157 ( 28.0%) |
| Likey Related | 2 ( 0.4%) | 2 ( 2.5%) | 4 ( 0.7%) |
| Not Related | 48 ( 10.0%) | 11 ( 13.8%) | 59 ( 10.5%) |
| Probably Related | 42 ( 8.8%) | 6 ( 7.5%) | 48 ( 8.6%) |
| Related | 3 ( 0.6%) | | 3 ( 0.5%) |
| Unlikely Related | 35 ( 7.3%) | 8 ( 10.0%) | 43 ( 7.7%) |
| Blood and lymphatic system disorders | 7 ( 1.5%) | 2 ( 2.5%) | 9 ( 1.6%) |
| Not Related | 4 ( 0.8%) | | 4 ( 0.7%) |
| Probably Related | 1 ( 0.2%) | 1 ( 1.3%) | 2 ( 0.4%) |
| Unlikely Related | 2 ( 0.4%) | 1 ( 1.3%) | 3 ( 0.5%) |
| Anemia | 1 ( 0.2%) | 1 ( 1.3%) | 2 ( 0.4%) |
| Probably Related | 1 ( 0.2%) | | 1 ( 0.2%) |
| Unlikely Related | | 1 ( 1.3%) | 1 ( 0.2%) |
| Hematuria | 1 ( 0.2%) | | 1 ( 0.2%) |
| Not Related | 1 ( 0.2%) | | 1 ( 0.2%) |
| Leukocytes | 6 ( 1.3%) | 2 ( 2.5%) | 8 ( 1.4%) |
| Not Related | 4 ( 0.8%) | | 4 ( 0.7%) |

*Created by D.D.G with SAS 9.4 on 11MAY2021*
*Program: SAS/folders/myfolders/NANOCOVAXP2/AE/UnSoliAE/AEREL2.SAS*

**Appendix 7: Unsolicited Adverse Events by System Organ Classes, Preferred Term and Relationship to Study Drug**

Safety Analysis Set- Placebo Vs. Nanocovax (25mcg/50mcg/75mcg

| Body System<br>Preferred Term<br>Relationship | Nanocovax<br>N=480 | Placebo<br>N= 80 | OVERALL<br>N=560 |
|---|---|---|---|
| Probably Related | | 1 ( 1.3%) | 1 ( 0.2%) |
| Unlikely Related | 2 ( 0.4%) | 1 ( 1.3%) | 3 ( 0.5%) |
| Cardiac disorders | 6 ( 1.3%) | 1 ( 1.3%) | 7 ( 1.3%) |
| Likey Related | 1 ( 0.2%) | | 1 ( 0.2%) |
| Not Related | 1 ( 0.2%) | 1 ( 1.3%) | 2 ( 0.4%) |
| Probably Related | 3 ( 0.6%) | | 3 ( 0.5%) |
| Unlikely Related | 1 ( 0.2%) | | 1 ( 0.2%) |
| Chest pain | 6 ( 1.3%) | | 6 ( 1.1%) |
| Likey Related | 1 ( 0.2%) | | 1 ( 0.2%) |
| Not Related | 1 ( 0.2%) | | 1 ( 0.2%) |
| Probably Related | 3 ( 0.6%) | | 3 ( 0.5%) |
| Unlikely Related | 1 ( 0.2%) | | 1 ( 0.2%) |
| Stable angina | | 1 ( 1.3%) | 1 ( 0.2%) |
| Not Related | | 1 ( 1.3%) | 1 ( 0.2%) |
| Tachycardia | 1 ( 0.2%) | | 1 ( 0.2%) |
| Probably Related | 1 ( 0.2%) | | 1 ( 0.2%) |

*Created by D.D.G with SAS 9.4 on 11MAY2021*
*Program: SAS/folders/myfolders/NANOCOVAXP2/AE/UnSoliAE/AEREL2.SAS*

**Appendix 7: Unsolicited Adverse Events by System Organ Classes, Preferred Term and Relationship to Study Drug**

Safety Analysis Set- Placebo Vs. Nanocovax (25mcg/50mcg/75mcg

| **Body System**<br>**Preferred Term**<br>**Relationship** | **Nanocovax**<br>**N=480** | **Placebo**<br>**N= 80** | **OVERALL**<br>**N=560** |
|---|---|---|---|
| Eye disorders | 4 ( 0.8%) | | 4 ( 0.7%) |
| Not Related | 3 ( 0.6%) | | 3 ( 0.5%) |
| Probably Related | 1 ( 0.2%) | | 1 ( 0.2%) |
| Conjunctivitis | 1 ( 0.2%) | | 1 ( 0.2%) |
| Not Related | 1 ( 0.2%) | | 1 ( 0.2%) |
| Eyes tearing | 1 ( 0.2%) | | 1 ( 0.2%) |
| Not Related | 1 ( 0.2%) | | 1 ( 0.2%) |
| Hordeolum | 1 ( 0.2%) | | 1 ( 0.2%) |
| Not Related | 1 ( 0.2%) | | 1 ( 0.2%) |
| Itching- eye area | 1 ( 0.2%) | | 1 ( 0.2%) |
| Probably Related | 1 ( 0.2%) | | 1 ( 0.2%) |
| Gastrointestinal disorders | 8 ( 1.7%) | 1 ( 1.3%) | 9 ( 1.6%) |
| Not Related | 4 ( 0.8%) | | 4 ( 0.7%) |
| Probably Related | 2 ( 0.4%) | | 2 ( 0.4%) |
| Unlikely Related | 2 ( 0.4%) | 1 ( 1.3%) | 3 ( 0.5%) |

*Created by D.D.G with SAS 9.4 on 11MAY2021*
*Program: SAS/folders/myfolders/NANOCOVAXP2/AE/UnSoliAE/AEREL2.SAS*

**Appendix 7: Unsolicited Adverse Events by System Organ Classes, Preferred Term and Relationship to Study Drug**

Safety Analysis Set- Placebo Vs. Nanocovax (25mcg/50mcg/75mcg

| Body System<br>Preferred Term<br>Relationship | Nanocovax<br>N=480 | Placebo<br>N= 80 | OVERALL<br>N=560 |
|---|---|---|---|
| Abdominal pains | 2 ( 0.4%) | | 2 ( 0.4%) |
| Probably Related | 1 ( 0.2%) | | 1 ( 0.2%) |
| Unlikely Related | 1 ( 0.2%) | | 1 ( 0.2%) |
| Diarhea | 1 ( 0.2%) | | 1 ( 0.2%) |
| Probably Related | 1 ( 0.2%) | | 1 ( 0.2%) |
| Dry mouth | 1 ( 0.2%) | | 1 ( 0.2%) |
| Probably Related | 1 ( 0.2%) | | 1 ( 0.2%) |
| Gastralgia | | 1 ( 1.3%) | 1 ( 0.2%) |
| Unlikely Related | | 1 ( 1.3%) | 1 ( 0.2%) |
| Gastritis | 1 ( 0.2%) | | 1 ( 0.2%) |
| Not Related | 1 ( 0.2%) | | 1 ( 0.2%) |
| Left upper quadrant pain | 1 ( 0.2%) | | 1 ( 0.2%) |
| Not Related | 1 ( 0.2%) | | 1 ( 0.2%) |
| Toothache | 3 ( 0.6%) | | 3 ( 0.5%) |
| Not Related | 2 ( 0.4%) | | 2 ( 0.4%) |

*Created by D.D.G with SAS 9.4 on 11MAY2021*
*Program: SAS/folders/myfolders/NANOCOVAXP2/AE/UnSoliAE/AEREL2.SAS*

**Appendix 7: Unsolicited Adverse Events by System Organ Classes, Preferred Term and Relationship to Study Drug**

Safety Analysis Set- Placebo Vs. Nanocovax (25mcg/50mcg/75mcg

| Body System<br>Preferred Term<br>Relationship | Nanocovax<br>N=480 | Placebo<br>N= 80 | OVERALL<br>N=560 |
|---|---|---|---|
| Unlikely Related | 1 ( 0.2%) | | 1 ( 0.2%) |
| General disorders and administration site conditions | 22 ( 4.6%) | 5 ( 6.3%) | 27 ( 4.8%) |
| Likey Related | | 1 ( 1.3%) | 1 ( 0.2%) |
| Not Related | 4 ( 0.8%) | 1 ( 1.3%) | 5 ( 0.9%) |
| Probably Related | 11 ( 2.3%) | 1 ( 1.3%) | 12 ( 2.1%) |
| Related | 2 ( 0.4%) | | 2 ( 0.4%) |
| Unlikely Related | 5 ( 1.0%) | 2 ( 2.5%) | 7 ( 1.3%) |
| Anorexia | 1 ( 0.2%) | | 1 ( 0.2%) |
| Probably Related | 1 ( 0.2%) | | 1 ( 0.2%) |
| Drowsy | | 1 ( 1.3%) | 1 ( 0.2%) |
| Likey Related | | 1 ( 1.3%) | 1 ( 0.2%) |
| Dyspepsia flatulence | 2 ( 0.4%) | | 2 ( 0.4%) |
| Probably Related | 2 ( 0.4%) | | 2 ( 0.4%) |
| Fatigue | 2 ( 0.4%) | 1 ( 1.3%) | 3 ( 0.5%) |
| Not Related | 1 ( 0.2%) | | 1 ( 0.2%) |
| Probably Related | 1 ( 0.2%) | | 1 ( 0.2%) |

*Created by D.D.G with SAS 9.4 on 11MAY2021*
*Program: SAS/folders/myfolders/NANOCOVAXP2/AE/UnSoliAE/AEREL2.SAS*

**Appendix 7: Unsolicited Adverse Events by System Organ Classes, Preferred Term and Relationship to Study Drug**

Safety Analysis Set- Placebo Vs. Nanocovax (25mcg/50mcg/75mcg

| Body System<br>Preferred Term<br>Relationship | Nanocovax<br>N=480 | Placebo<br>N= 80 | OVERALL<br>N=560 |
|---|---|---|---|
| Unlikely Related | | 1 ( 1.3%) | 1 ( 0.2%) |
| Fever | 5 ( 1.0%) | 1 ( 1.3%) | 6 ( 1.1%) |
| Not Related | 1 ( 0.2%) | | 1 ( 0.2%) |
| Probably Related | 1 ( 0.2%) | | 1 ( 0.2%) |
| Related | 1 ( 0.2%) | | 1 ( 0.2%) |
| Unlikely Related | 2 ( 0.4%) | 1 ( 1.3%) | 3 ( 0.5%) |
| Headache | 1 ( 0.2%) | | 1 ( 0.2%) |
| Not Related | 1 ( 0.2%) | | 1 ( 0.2%) |
| Hypothermia | 10 ( 2.1%) | 2 ( 2.5%) | 12 ( 2.1%) |
| Not Related | 1 ( 0.2%) | 1 ( 1.3%) | 2 ( 0.4%) |
| Probably Related | 6 ( 1.3%) | 1 ( 1.3%) | 7 ( 1.3%) |
| Unlikely Related | 3 ( 0.6%) | | 3 ( 0.5%) |
| Insomnia | 2 ( 0.4%) | | 2 ( 0.4%) |
| Not Related | 1 ( 0.2%) | | 1 ( 0.2%) |
| Probably Related | 1 ( 0.2%) | | 1 ( 0.2%) |
| Loss of taste | 1 ( 0.2%) | | 1 ( 0.2%) |

*Created by D.D.G with SAS 9.4 on 11MAY2021*
*Program: SAS/folders/myfolders/NANOCOVAXP2/AE/UnSoliAE/AEREL2.SAS*

**Appendix 7: Unsolicited Adverse Events by System Organ Classes, Preferred Term and Relationship to Study Drug**

Safety Analysis Set- Placebo Vs. Nanocovax (25mcg/50mcg/75mcg

| Body System<br>Preferred Term<br>Relationship | Nanocovax<br>N=480 | Placebo<br>N= 80 | OVERALL<br>N=560 |
|---|---|---|---|
| Probably Related | 1 ( 0.2%) | | 1 ( 0.2%) |
| Tenderness | 1 ( 0.2%) | | 1 ( 0.2%) |
| Related | 1 ( 0.2%) | | 1 ( 0.2%) |
| Immune system disorders | 3 ( 0.6%) | | 3 ( 0.5%) |
| Not Related | 2 ( 0.4%) | | 2 ( 0.4%) |
| Probably Related | 1 ( 0.2%) | | 1 ( 0.2%) |
| Allergic reaction | 2 ( 0.4%) | | 2 ( 0.4%) |
| Not Related | 1 ( 0.2%) | | 1 ( 0.2%) |
| Probably Related | 1 ( 0.2%) | | 1 ( 0.2%) |
| Anaphylaxis- Grade I | 1 ( 0.2%) | | 1 ( 0.2%) |
| Not Related | 1 ( 0.2%) | | 1 ( 0.2%) |
| Infections and infestations | 3 ( 0.6%) | 1 ( 1.3%) | 4 ( 0.7%) |
| Not Related | 2 ( 0.4%) | 1 ( 1.3%) | 3 ( 0.5%) |
| Probably Related | 1 ( 0.2%) | | 1 ( 0.2%) |
| Atrioventricular node abscess | 1 ( 0.2%) | | 1 ( 0.2%) |

*Created by D.D.G with SAS 9.4 on 11MAY2021*
*Program: SAS/folders/myfolders/NANOCOVAXP2/AE/UnSoliAE/AEREL2.SAS*

**Appendix 7: Unsolicited Adverse Events by System Organ Classes, Preferred Term and Relationship to Study Drug**

Safety Analysis Set- Placebo Vs. Nanocovax (25mcg/50mcg/75mcg

| Body System<br>Preferred Term<br>Relationship | Nanocovax<br>N=480 | Placebo<br>N= 80 | OVERALL<br>N=560 |
|---|---|---|---|
| Not Related | 1 ( 0.2%) | | 1 ( 0.2%) |
| Flu | 1 ( 0.2%) | 1 ( 1.3%) | 2 ( 0.4%) |
| Not Related | | 1 ( 1.3%) | 1 ( 0.2%) |
| Probably Related | 1 ( 0.2%) | | 1 ( 0.2%) |
| Sepsis | 1 ( 0.2%) | | 1 ( 0.2%) |
| Not Related | 1 ( 0.2%) | | 1 ( 0.2%) |
| Injury, poisoning and procedural complications | 1 ( 0.2%) | | 1 ( 0.2%) |
| Not Related | 1 ( 0.2%) | | 1 ( 0.2%) |
| Broken Bone, Right Heel | 1 ( 0.2%) | | 1 ( 0.2%) |
| Not Related | 1 ( 0.2%) | | 1 ( 0.2%) |
| Investigations | 6 ( 1.3%) | 3 ( 3.8%) | 9 ( 1.6%) |
| Probably Related | 3 ( 0.6%) | 2 ( 2.5%) | 5 ( 0.9%) |
| Unlikely Related | 3 ( 0.6%) | 1 ( 1.3%) | 4 ( 0.7%) |
| Elevated White Blood Cells | 5 ( 1.0%) | 1 ( 1.3%) | 6 ( 1.1%) |
| Probably Related | 2 ( 0.4%) | 1 ( 1.3%) | 3 ( 0.5%) |

*Created by D.D.G with SAS 9.4 on 11MAY2021*
*Program: SAS/folders/myfolders/NANOCOVAXP2/AE/UnSoliAE/AEREL2.SAS*

**Appendix 7: Unsolicited Adverse Events by System Organ Classes, Preferred Term and Relationship to Study Drug**

Safety Analysis Set- Placebo Vs. Nanocovax (25mcg/50mcg/75mcg

| Body System<br>Preferred Term<br>Relationship | Nanocovax<br>N=480 | Placebo<br>N= 80 | OVERALL<br>N=560 |
|---|---|---|---|
| Unlikely Related | 3 ( 0.6%) | | 3 ( 0.5%) |
| Elevated liver function tests | | 2 ( 2.5%) | 2 ( 0.4%) |
| Probably Related | | 1 ( 1.3%) | 1 ( 0.2%) |
| Unlikely Related | | 1 ( 1.3%) | 1 ( 0.2%) |
| WBC formula shifted | 1 ( 0.2%) | | 1 ( 0.2%) |
| Probably Related | 1 ( 0.2%) | | 1 ( 0.2%) |
| Metabolism and nutrition disorders | 11 ( 2.3%) | 2 ( 2.5%) | 13 ( 2.3%) |
| Not Related | 7 ( 1.5%) | 1 ( 1.3%) | 8 ( 1.4%) |
| Unlikely Related | 4 ( 0.8%) | 1 ( 1.3%) | 5 ( 0.9%) |
| Hyperglycemia | 11 ( 2.3%) | 2 ( 2.5%) | 13 ( 2.3%) |
| Not Related | 6 ( 1.3%) | 1 ( 1.3%) | 7 ( 1.3%) |
| Probably Related | 1 ( 0.2%) | | 1 ( 0.2%) |
| Unlikely Related | 4 ( 0.8%) | 1 ( 1.3%) | 5 ( 0.9%) |
| Urinary glucose | 5 ( 1.0%) | 1 ( 1.3%) | 6 ( 1.1%) |
| Not Related | 4 ( 0.8%) | 1 ( 1.3%) | 5 ( 0.9%) |
| Unlikely Related | 1 ( 0.2%) | | 1 ( 0.2%) |

**Appendix 7: Unsolicited Adverse Events by System Organ Classes, Preferred Term and Relationship to Study Drug**

Safety Analysis Set- Placebo Vs. Nanocovax (25mcg/50mcg/75mcg

| Body System<br>Preferred Term<br>Relationship | Nanocovax<br>N=480 | Placebo<br>N= 80 | OVERALL<br>N=560 |
|---|---|---|---|
| Musculoskeletal and connective tissue disorders | 21 ( 4.4%) | 11 ( 13.8%) | 32 ( 5.7%) |
| Likey Related | 1 ( 0.2%) | | 1 ( 0.2%) |
| Not Related | 10 ( 2.1%) | 7 ( 8.8%) | 17 ( 3.0%) |
| Probably Related | 6 ( 1.3%) | | 6 ( 1.1%) |
| Related | 2 ( 0.4%) | | 2 ( 0.4%) |
| Unlikely Related | 2 ( 0.4%) | 4 ( 5.0%) | 6 ( 1.1%) |
| 2-sided breast pain | 1 ( 0.2%) | | 1 ( 0.2%) |
| Probably Related | 1 ( 0.2%) | | 1 ( 0.2%) |
| Arthralgia | 6 ( 1.3%) | 3 ( 3.8%) | 9 ( 1.6%) |
| Not Related | 2 ( 0.4%) | 1 ( 1.3%) | 3 ( 0.5%) |
| Probably Related | 2 ( 0.4%) | | 2 ( 0.4%) |
| Unlikely Related | 2 ( 0.4%) | 2 ( 2.5%) | 4 ( 0.7%) |
| Arthritis | | 2 ( 2.5%) | 2 ( 0.4%) |
| Not Related | | 1 ( 1.3%) | 1 ( 0.2%) |
| Unlikely Related | | 1 ( 1.3%) | 1 ( 0.2%) |
| Back pain | 2 ( 0.4%) | 1 ( 1.3%) | 3 ( 0.5%) |

**Appendix 7: Unsolicited Adverse Events by System Organ Classes, Preferred Term and Relationship to Study Drug**

Safety Analysis Set- Placebo Vs. Nanocovax (25mcg/50mcg/75mcg

| Body System<br>Preferred Term<br>Relationship | Nanocovax<br>N=480 | Placebo<br>N= 80 | OVERALL<br>N=560 |
|---|---|---|---|
| Not Related | 2 ( 0.4%) | 1 ( 1.3%) | 3 ( 0.5%) |
| Cervical scapulohumeral syndrome | 1 ( 0.2%) | | 1 ( 0.2%) |
| Probably Related | 1 ( 0.2%) | | 1 ( 0.2%) |
| Degenerative Spine | 2 ( 0.4%) | | 2 ( 0.4%) |
| Not Related | 2 ( 0.4%) | | 2 ( 0.4%) |
| Disc herniation | 1 ( 0.2%) | | 1 ( 0.2%) |
| Likey Related | 1 ( 0.2%) | | 1 ( 0.2%) |
| Hand pain | 2 ( 0.4%) | 1 ( 1.3%) | 3 ( 0.5%) |
| Not Related | 1 ( 0.2%) | 1 ( 1.3%) | 2 ( 0.4%) |
| Probably Related | 1 ( 0.2%) | | 1 ( 0.2%) |
| Knee pain | 1 ( 0.2%) | 1 ( 1.3%) | 2 ( 0.4%) |
| Not Related | 1 ( 0.2%) | 1 ( 1.3%) | 2 ( 0.4%) |
| Myalgia | 5 ( 1.0%) | | 5 ( 0.9%) |
| Not Related | 1 ( 0.2%) | | 1 ( 0.2%) |
| Probably Related | 1 ( 0.2%) | | 1 ( 0.2%) |

*Created by D.D.G with SAS 9.4 on 11MAY2021*
*Program: SAS/folders/myfolders/NANOCOVAXP2/AE/UnSoliAE/AEREL2.SAS*

**Appendix 7: Unsolicited Adverse Events by System Organ Classes, Preferred Term and Relationship to Study Drug**

Safety Analysis Set- Placebo Vs. Nanocovax (25mcg/50mcg/75mcg

| Body System<br>Preferred Term<br>Relationship | Nanocovax<br>N=480 | Placebo<br>N= 80 | OVERALL<br>N=560 |
|---|---|---|---|
| Related | 2 ( 0.4%) | | 2 ( 0.4%) |
| Unlikely Related | 1 ( 0.2%) | | 1 ( 0.2%) |
| Neck shoulder pain | 1 ( 0.2%) | 3 ( 3.8%) | 4 ( 0.7%) |
| Not Related | | 1 ( 1.3%) | 1 ( 0.2%) |
| Probably Related | | 1 ( 1.3%) | 1 ( 0.2%) |
| Unlikely Related | 1 ( 0.2%) | 1 ( 1.3%) | 2 ( 0.4%) |
| Osteoarthritis | 1 ( 0.2%) | | 1 ( 0.2%) |
| Not Related | 1 ( 0.2%) | | 1 ( 0.2%) |
| Rheumatoid arthritis | 1 ( 0.2%) | 1 ( 1.3%) | 2 ( 0.4%) |
| Not Related | 1 ( 0.2%) | | 1 ( 0.2%) |
| Unlikely Related | | 1 ( 1.3%) | 1 ( 0.2%) |
| Sore feet | | 1 ( 1.3%) | 1 ( 0.2%) |
| Not Related | | 1 ( 1.3%) | 1 ( 0.2%) |
| Spinal muscle pain | 1 ( 0.2%) | | 1 ( 0.2%) |
| Not Related | 1 ( 0.2%) | | 1 ( 0.2%) |

*Created by D.D.G with SAS 9.4 on 11MAY2021*
*Program: SAS/folders/myfolders/NANOCOVAXP2/AE/UnSoliAE/AEREL2.SAS*

**Appendix 7: Unsolicited Adverse Events by System Organ Classes, Preferred Term and Relationship to Study Drug**

Safety Analysis Set- Placebo Vs. Nanocovax (25mcg/50mcg/75mcg

| **Body System<br>Preferred Term<br>Relationship** | **Nanocovax<br>N=480** | **Placebo<br>N= 80** | **OVERALL<br>N=560** |
|---|---|---|---|
| Spinal pain | 2 ( 0.4%) | | 2 ( 0.4%) |
| Probably Related | 2 ( 0.4%) | | 2 ( 0.4%) |
| Nervous system disorders | 7 ( 1.5%) | 3 ( 3.8%) | 10 ( 1.8%) |
| Not Related | 1 ( 0.2%) | | 1 ( 0.2%) |
| Probably Related | 3 ( 0.6%) | 1 ( 1.3%) | 4 ( 0.7%) |
| Unlikely Related | 3 ( 0.6%) | 2 ( 2.5%) | 5 ( 0.9%) |
| Dizziness | 4 ( 0.8%) | 2 ( 2.5%) | 6 ( 1.1%) |
| Not Related | 1 ( 0.2%) | | 1 ( 0.2%) |
| Probably Related | 1 ( 0.2%) | 1 ( 1.3%) | 2 ( 0.4%) |
| Unlikely Related | 2 ( 0.4%) | 1 ( 1.3%) | 3 ( 0.5%) |
| Headache | 5 ( 1.0%) | | 5 ( 0.9%) |
| Not Related | 1 ( 0.2%) | | 1 ( 0.2%) |
| Probably Related | 3 ( 0.6%) | | 3 ( 0.5%) |
| Unlikely Related | 1 ( 0.2%) | | 1 ( 0.2%) |
| Vestibular disorders | | 1 ( 1.3%) | 1 ( 0.2%) |
| Unlikely Related | | 1 ( 1.3%) | 1 ( 0.2%) |

*Created by D.D.G with SAS 9.4 on 11MAY2021*
*Program: SAS/folders/myfolders/NANOCOVAXP2/AE/UnSoliAE/AEREL2.SAS*

**Appendix 7: Unsolicited Adverse Events by System Organ Classes, Preferred Term and Relationship to Study Drug**

Safety Analysis Set- Placebo Vs. Nanocovax (25mcg/50mcg/75mcg

| **Body System**<br>**Preferred Term**<br>**Relationship** | **Nanocovax**<br>**N=480** | **Placebo**<br>**N= 80** | **OVERALL**<br>**N=560** |
|---|---|---|---|
| Renal and urinary disorders | 12 ( 2.5%) | 2 ( 2.5%) | 14 ( 2.5%) |
| Not Related | 6 ( 1.3%) | | 6 ( 1.1%) |
| Probably Related | 1 ( 0.2%) | 1 ( 1.3%) | 2 ( 0.4%) |
| Unlikely Related | 5 ( 1.0%) | 1 ( 1.3%) | 6 ( 1.1%) |
| Cystitis | 1 ( 0.2%) | | 1 ( 0.2%) |
| Unlikely Related | 1 ( 0.2%) | | 1 ( 0.2%) |
| Hematuria | 7 ( 1.5%) | 2 ( 2.5%) | 9 ( 1.6%) |
| Not Related | 3 ( 0.6%) | | 3 ( 0.5%) |
| Probably Related | | 1 ( 1.3%) | 1 ( 0.2%) |
| Unlikely Related | 4 ( 0.8%) | 1 ( 1.3%) | 5 ( 0.9%) |
| Painful urination | 1 ( 0.2%) | | 1 ( 0.2%) |
| Probably Related | 1 ( 0.2%) | | 1 ( 0.2%) |
| Urinary tract infection | 3 ( 0.6%) | | 3 ( 0.5%) |
| Not Related | 3 ( 0.6%) | | 3 ( 0.5%) |
| Reproductive system and breast disorders | 2 ( 0.4%) | 2 ( 2.5%) | 4 ( 0.7%) |
| Not Related | 1 ( 0.2%) | | 1 ( 0.2%) |

*Created by D.D.G with SAS 9.4 on 11MAY2021*
*Program: SAS/folders/myfolders/NANOCOVAXP2/AE/UnSoliAE/AEREL2.SAS*

**Appendix 7: Unsolicited Adverse Events by System Organ Classes, Preferred Term and Relationship to Study Drug**

Safety Analysis Set- Placebo Vs. Nanocovax (25mcg/50mcg/75mcg

| Body System<br>Preferred Term<br>Relationship | Nanocovax<br>N=480 | Placebo<br>N= 80 | OVERALL<br>N=560 |
|---|---|---|---|
| Probably Related | 1 ( 0.2%) | 2 ( 2.5%) | 3 ( 0.5%) |
| Menstruation irregular | 2 ( 0.4%) | 2 ( 2.5%) | 4 ( 0.7%) |
| Not Related | 1 ( 0.2%) | | 1 ( 0.2%) |
| Probably Related | 1 ( 0.2%) | 2 ( 2.5%) | 3 ( 0.5%) |
| Respiratory, thoracic and mediastinal disorders | 43 ( 9.0%) | 6 ( 7.5%) | 49 ( 8.8%) |
| Likey Related | 1 ( 0.2%) | | 1 ( 0.2%) |
| Not Related | 12 ( 2.5%) | 2 ( 2.5%) | 14 ( 2.5%) |
| Probably Related | 14 ( 2.9%) | 3 ( 3.8%) | 17 ( 3.0%) |
| Unlikely Related | 16 ( 3.3%) | 1 ( 1.3%) | 17 ( 3.0%) |
| Acute sinusitis | 1 ( 0.2%) | | 1 ( 0.2%) |
| Unlikely Related | 1 ( 0.2%) | | 1 ( 0.2%) |
| Allergic Nasopharyngitis | 1 ( 0.2%) | | 1 ( 0.2%) |
| Not Related | 1 ( 0.2%) | | 1 ( 0.2%) |
| Allergic rhinitis | 1 ( 0.2%) | | 1 ( 0.2%) |
| Probably Related | 1 ( 0.2%) | | 1 ( 0.2%) |

*Created by D.D.G with SAS 9.4 on 11MAY2021*
*Program: SAS/folders/myfolders/NANOCOVAXP2/AE/UnSoliAE/AEREL2.SAS*

**Appendix 7: Unsolicited Adverse Events by System Organ Classes, Preferred Term and Relationship to Study Drug**

Safety Analysis Set- Placebo Vs. Nanocovax (25mcg/50mcg/75mcg

| Body System<br>Preferred Term<br>Relationship | Nanocovax<br>N=480 | Placebo<br>N= 80 | OVERALL<br>N=560 |
|---|---|---|---|
| Amidan swelling | 1 ( 0.2%) | | 1 ( 0.2%) |
| Not Related | 1 ( 0.2%) | | 1 ( 0.2%) |
| Cough | 9 ( 1.9%) | 2 ( 2.5%) | 11 ( 2.0%) |
| Not Related | 2 ( 0.4%) | | 2 ( 0.4%) |
| Probably Related | 6 ( 1.3%) | 1 ( 1.3%) | 7 ( 1.3%) |
| Unlikely Related | 1 ( 0.2%) | 1 ( 1.3%) | 2 ( 0.4%) |
| Dyspnea | 2 ( 0.4%) | | 2 ( 0.4%) |
| Not Related | 1 ( 0.2%) | | 1 ( 0.2%) |
| Unlikely Related | 1 ( 0.2%) | | 1 ( 0.2%) |
| Hoarseness | 1 ( 0.2%) | | 1 ( 0.2%) |
| Probably Related | 1 ( 0.2%) | | 1 ( 0.2%) |
| Nasal Discharge | 6 ( 1.3%) | 2 ( 2.5%) | 8 ( 1.4%) |
| Not Related | 1 ( 0.2%) | 1 ( 1.3%) | 2 ( 0.4%) |
| Probably Related | 1 ( 0.2%) | | 1 ( 0.2%) |
| Unlikely Related | 4 ( 0.8%) | 1 ( 1.3%) | 5 ( 0.9%) |
| Nasal pain | | 1 ( 1.3%) | 1 ( 0.2%) |

*Created by D.D.G with SAS 9.4 on 11MAY2021*
*Program: SAS/folders/myfolders/NANOCOVAXP2/AE/UnSoliAE/AEREL2.SAS*

**Appendix 7: Unsolicited Adverse Events by System Organ Classes, Preferred Term and Relationship to Study Drug**

Safety Analysis Set- Placebo Vs. Nanocovax (25mcg/50mcg/75mcg

| Body System<br>Preferred Term<br>Relationship | Nanocovax<br>N=480 | Placebo<br>N= 80 | OVERALL<br>N=560 |
|---|---|---|---|
| Not Related | | 1 ( 1.3%) | 1 ( 0.2%) |
| Sinusitis | 1 ( 0.2%) | | 1 ( 0.2%) |
| Unlikely Related | 1 ( 0.2%) | | 1 ( 0.2%) |
| Sore Throat | 23 ( 4.8%) | 4 ( 5.0%) | 27 ( 4.8%) |
| Likey Related | 1 ( 0.2%) | | 1 ( 0.2%) |
| Not Related | 7 ( 1.5%) | 1 ( 1.3%) | 8 ( 1.4%) |
| Probably Related | 9 ( 1.9%) | 2 ( 2.5%) | 11 ( 2.0%) |
| Unlikely Related | 6 ( 1.3%) | 1 ( 1.3%) | 7 ( 1.3%) |
| Throat sensation disorders | 2 ( 0.4%) | | 2 ( 0.4%) |
| Unlikely Related | 2 ( 0.4%) | | 2 ( 0.4%) |
| Upper respiratory tract infection | 1 ( 0.2%) | | 1 ( 0.2%) |
| Not Related | 1 ( 0.2%) | | 1 ( 0.2%) |
| Skin and subcutaneous tissue disorders | 6 ( 1.3%) | 2 ( 2.5%) | 8 ( 1.4%) |
| Likey Related | | 1 ( 1.3%) | 1 ( 0.2%) |
| Not Related | 1 ( 0.2%) | 1 ( 1.3%) | 2 ( 0.4%) |
| Probably Related | 5 ( 1.0%) | | 5 ( 0.9%) |

**Appendix 7: Unsolicited Adverse Events by System Organ Classes, Preferred Term and Relationship to Study Drug**

Safety Analysis Set- Placebo Vs. Nanocovax (25mcg/50mcg/75mcg

| Body System<br>Preferred Term<br>Relationship | Nanocovax<br>N=480 | Placebo<br>N= 80 | OVERALL<br>N=560 |
|---|---|---|---|
| Burn | 1 ( 0.2%) | | 1 ( 0.2%) |
| Not Related | 1 ( 0.2%) | | 1 ( 0.2%) |
| Itching- head skin | | 1 ( 1.3%) | 1 ( 0.2%) |
| Not Related | | 1 ( 1.3%) | 1 ( 0.2%) |
| Itchy rash | 3 ( 0.6%) | 1 ( 1.3%) | 4 ( 0.7%) |
| Likey Related | | 1 ( 1.3%) | 1 ( 0.2%) |
| Probably Related | 3 ( 0.6%) | | 3 ( 0.5%) |
| Lumps at the injection site | 1 ( 0.2%) | | 1 ( 0.2%) |
| Probably Related | 1 ( 0.2%) | | 1 ( 0.2%) |
| Skin infection | 1 ( 0.2%) | | 1 ( 0.2%) |
| Probably Related | 1 ( 0.2%) | | 1 ( 0.2%) |
| Vascular disorders | 6 ( 1.3%) | 2 ( 2.5%) | 8 ( 1.4%) |
| Not Related | 3 ( 0.6%) | 2 ( 2.5%) | 5 ( 0.9%) |
| Probably Related | 3 ( 0.6%) | | 3 ( 0.5%) |

**Appendix 7: Unsolicited Adverse Events by System Organ Classes, Preferred Term and Relationship to Study Drug**

Safety Analysis Set- Placebo Vs. Nanocovax (25mcg/50mcg/75mcg

| **Body System**<br>**Preferred Term**<br>**Relationship** | **Nanocovax**<br>**N=480** | **Placebo**<br>**N= 80** | **OVERALL**<br>**N=560** |
|---|---|---|---|
| Hypertension | 6 ( 1.3%) | 1 ( 1.3%) | 7 ( 1.3%) |
| Not Related | 3 ( 0.6%) | 1 ( 1.3%) | 4 ( 0.7%) |
| Probably Related | 3 ( 0.6%) | | 3 ( 0.5%) |
| Hypotension | | 1 ( 1.3%) | 1 ( 0.2%) |
| Not Related | | 1 ( 1.3%) | 1 ( 0.2%) |

*Created by D.D.G with SAS 9.4 on 11MAY2021*
*Program: SAS/folders/myfolders/NANOCOVAXP2/AE/UnSoliAE/AEREL2.SAS*
